# Reports of deaths are an exaggeration: German (PCR-test-positive) fatality counts during the SARS-CoV-2 era in the context of all-cause mortality

**DOI:** 10.1101/2022.11.18.22282534

**Authors:** Robert Rockenfeller, Michael Günther, Falk Mörl

## Abstract

As of March 2020, counts of SARS-CoV-2-related (‘CoViD-19’) deaths have been key numbers for justifying severe political, social, and economical measures put into action by authorities world-wide. A particular focus thereby was the concomitant excess mortality (EM), i.e. fatalities above the normally expected all-cause mortality (AM). Recent reports and studies, inter alia by the WHO, estimated the SARS-CoV-2-related EM in Germany between 2020 and 2021 as high as 200,000. In this study, we attempt to scrutinise these numbers by putting them into the context of German AM since the year 2000.

To this end, we propose two straightforward, low-parametric models to estimate German AM, and thus EM, for the years 2020 and 2021 as well as the flu seasons 2020/21 and 2021/22. Additionally, we give a forecast of the AM expected in 2022. After having derived age-cohort-specific mortality rates out of historical data, weighted with their corresponding demographic proportion, EM is obtained by subtracting (model-)calculated AM counts from observed ones. For Germany, we find even an overall negative EM (‘under-mortality’) of about -18,500 for the year 2020, and a minor positive EM of about 7,000 for 2021, unveiling that officially communicated EM numbers are a great exaggeration. Further, putting CoViD-19 “cases” (defined by positive PCR test outcomes) and their related death counts into the context of AM, we are able to estimate how many Germans have died *due to* rather than *with* CoViD-19; an analysis not provided by the appropriate authority, the RKI. Thereby, varying governmental PCR test strategies are shown to heavily obscure reliable estimations of SARS-CoV-2-related EM, particularly within the second year of the proclaimed pandemic.

## 1. Introduction

A *mortality rate* is the number (*count*) of how many people have died in a specific country within a defined time interval, *normalised* to the whole number of inhabitants (population size). Counts may also be done, for example, only within age cohorts, and the corresponding rates are then usually normalised to the cohort size. If counts do not ask for any death condition then all-cause mortality (AM) is registered. The balance of *all-cause* mortality rate (AMR) and birth rate determines whether there is overall growth or loss in the population. Across the world, the most usual and wide-spread way of documenting an all-cause mortality count (AMC) and an AMR by a state authority is to report them annually. Depending on the geographical, climatic, and socio-economic situation, AMRs differ between countries [1]. In Europe and in the United States of America, for example, the annual AMRs are a little bit higher than one percent per year. Unsurprisingly, the AMRs in the European and North-American societies are mainly determined by the high probabilities of the eldest passing away [2]. The world-wide AMR has been generally decreasing during the last hundred years (see e.g. [3, 4, 5, 6]).

From the nineteenth century until now, the number of deaths due to infectious diseases were documented in more developed countries. Infectious diseases have a distinguishable impact on mortality. In Spain, from 1980 until 2011 as an example, mortality related to infectious diseases accounted for almost 15% of the AM [7], even though this percentage was decreasing during that period. As another example, data from cemeteries document a cholera epidemic in Brazil from 1855 to 1856, which resulted in as much as 70% more death counts in slaves than in free people, whereas the disease-specific mortality rate was identical [8]. Furthermore, people of different origin showed different mortality rates.

The influenza pandemic between 1918 and 1921 significantly increased the mortality in European countries [9] as well as in Arizona [10]. During that period, increased mortality occurred within few weeks, and mainly people between 20 and 50 years of age were affected by this influenza pandemic. A socio-economic phenomenon could be identified in Estonia after the year 1989, where the mortality in poorly educated subjects increased, while well-educated subjects had a constant mortality [11]. Taking the 2010-2011 Cholera epidemic in Haiti [12] as an example for the dynamics of infectious diseases, the AMR increased by about a factor of almost three over ten weeks; after that, the AMR decreased to values lower than usual. The disease-related mortality rates differed regionally.

In summary, a typical epidemic mortality pattern can be observed: initially, the mortality increases by a factor of up to ten within few weeks; after this period, there is usually mortality below average for several weeks. Accounting mortality over a whole year in extreme situations like the influenza pandemic one hundred years ago, the annual AMR can be as high as two thirds [9, 10] above the regular values (an annual excess of 66%). As a rule however, the all-cause and all-age mortality is dominated by specific disease sensitivities of some cohorts within the population, which may be old people or subjects with specific (health) conditions [2, 10]. Moreover, socio-economic factors can have a strong longer-term impact on mortality rates [11].

When proclaiming a SARS-CoV-2 pandemic in Europe in March 2020, the leading politicians of almost all northern, central, western, and southern European countries argued the same way, namely, that CoViD-19 (or short: ‘C19’) would presumably kill more people than usual. In other words: A significant increase in mortality was to be expected. Even in late May 2021 [13], German media and government officials in unison referred to weekly 1,300 deaths related to CoViD-19. This would have indeed been an excess mortality (EM) of 6.7% in Germany, given that, in 2021, about 19,550 fatalities per week were the regular mortality background (according to our model-based calculation of expected AM, see below in this paper), i.e. AM without an ongoing pandemic. However, summing daily AMCs according to the data collected by ‘Statistisches Bundesamt’ (Destatis; the German bureau for statistics) [14] resulted in exactly 18,579 persons who had died in Germany in the calendar week (#19) just before the above cited press conference. Thus, the “awfully high number” of 1,300 ‘CoViD-19 deaths’, which were announced with great media impetus by both the German minister of health and the head of the ‘Robert-Koch-Institut’ (RKI; the German centre for disease control), occurred in a week showing mortality *below* expectations (−5%). This ‘under-mortality’ has nothing of a deadly epidemic scenario at all. In any case, we had already wondered in the earliest ‘pandemic’ days, and nowadays even much more, where the vast majority of the media, in unison with authorities and their scientific consultants, took such large numbers from. More explicitly, we wondered why the German authorities refused to provide context to the public in order to assess the obtrusively proclaimed number of SARS-CoV-2-related (‘CoViD-19’) deaths. For the commonality, these numbers looked like (significant) excess mortality.

In March 2022, the World Health Organisation (WHO) published their latest update of SARS-CoV-2-related (“associated with COVID-19”) excess mortality in Germany during the years 2020 and 2021 [15], with a two-years total of 194,988. They had calculated these numbers using a Bayesian sampling model with spline-based seasonal variation [16, 17]. Therewith, the C19 deaths counted by German authorities had even been claimed to under-estimate the actual EM. Later, a WHO-endorsed model upgrade [18] provided a reduction to a two-years total of 122,000 excess deaths, i.e. close to the German authorities’ C19 death counts. Anyway, the currently available WHO data set [15] still contains the excess number 194,988.

With using only such data for analysis that have been published by German authorities themselves, we now aim in this study at providing an estimate of all-cause and SARS-CoV-2-related mortality, including excess numbers, for a well-founded, sound, and reliable final assessment of the epidemiological impact of CoViD-19 in Germany between April 2020 and late summer 2022. Particularly, we provide some missing context for all-cause and SARS-CoV-2-related as well as excess mortality numbers, annually and flu-seasonally.

## 2. Methods

### 2.1. Terminology and mathematical symbols

We start our considerations with a synopsis of the terminology used, and some explanations that also include abbreviations as well as mathematical symbols introduced.

A “case” of SARS-CoV-2 infection is indicated per legal definition, by a person being tested positively using a PCR test [19] (although strong indications of not being infection-indicative at all, see e.g. [20]), which is applied to sputum taken from the person’s throat or fluid from the nasopharynx. The German authorities, like most authorities around the world, relate a death to SARS-CoV-2, if the deceased had received such a positive PCR test result and (presumably, since criteria are not transparently communicated to the public, at least in Germany) had shown some clinical symptoms of CoViD-19 before passing away. We use the term *C19 death* as an abbreviation of such a putatively SARS-CoV-2-related, PCR-test-conditional (CoViD-19) death.

In the following brief overview, we introduce all essential abbreviations, despite some doubling with the introduction:

- AM for ‘all-cause mortality’, i.e. deaths occurring *due to any cause*.
- AMC for an ‘AM count’, i.e. the sum of registered cases of deceased persons within a given time interval (i.e. *per time unit*), whether a day, week, year, or season; mathematical symbol for any AMC: *D*_*AM*_ or, if referring to a particular cohort, *D*_*AM,coh*_.
- AMR for ‘AM rate’, i.e. the ratio of an AMC within a group (e.g. age cohort or specific population) to the sum of all group members (e.g. cohort size *N*_*coh*_ or number of inhabitants *N*_*pop*_); mathematical symbol for any AMR: *r*_*AM*_.
- PF for ‘PCR-test-conditional fatality’, i.e. deaths occurring *with* the deceased persons having received a positive PCR test result some time before passing away; in the following, ‘PCR-test-conditional’ is further contracted to ‘PCR-conditional’.
- PFC for a ‘PF count’, i.e. the sum of registered cases of PCR-conditionally deceased persons within a given time interval; mathematical symbol for any PFC: *D*_*PF*_.
- PFR for ‘PF rate’, i.e. the ratio of a PFC within a group (e.g. age cohort or specific population) to the sum of all group members (e.g. *PCR*_*pos*_); mathematical symbol for any PFR: *r*_*PF*_.
- EM for ‘excess mortality’, i.e. observed mortality (whether all-cause or any conditional) beyond expected mortality (usually, like in this study, AM).
- EMC for ‘EM count’, i.e. the difference between an observed mortality count (in this study, AMC or PFC) and the expected AMC.
- SMR for ‘standardised mortality ratio’, i.e. the ratio, within a group of persons fulfilling specific conditions (e.g. PCR-positive or with a disease), of an observed mortality count (in this study, PFC) and the expected one (usually, like in this study, AMC); mathematical symbol for the SMR of PCR-positive persons: 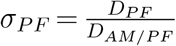, with *D*_*AM*|*PF*_ being the number of all-cause deaths *expected among the group members*.
- The mathematical symbol *n*_*tpp*_ represents the number of PCR *tests per person* in a given time interval.

### 2.2. Consulted data sets

Here is a concise synopsis of the German data sources that we based the present analysis and model development on:

- Earlier (2000-2015) all-cause daily and weekly death counts (broken down by age cohorts) were taken from the Destatis webpage at [21].
- Later (2016-2021) all-cause daily and weekly death counts (broken down by age cohorts) were taken from the Destatis webpage at [14].
- Weekly C19 death counts (broken down by age cohorts) were taken from the RKI webpage at [22].
- Counts of weekly SARS-CoV-2 “cases” (i.e. numbers of positive PCR tests, broken down by age cohorts; for person counts, we relate to Sec. 2.5 and Appendix A) were taken from the RKI webpage at [23].
- The demographic (age cohort) distribution until 2020 (based on the 2011 census) and its presumable distribution onwards (based on the most plausible scenario, the default model variant V1) were taken from the Destatis webpage at [24].
- Estimated counts of seasonal flu deaths were taken from the 2018 influenza report by the RKI: [25].

To make all above data sets comparable, weekly time resolution was chosen as a default for both death counts and putative SARS-CoV-2 “cases” (in fact, counts of PCR-positive persons), and thus for quantifying AMCs and PFCs (i.e. counts per week). For allowing specific comparisons or statements, data were occasionally contracted to annual values, and to address and solve some subtle modelling issues, we sometimes went back to daily resolution, which is then pointed out explicitly. The age cohorts were categorised in eight groups in terms of the unit *year* : 0-29, 30-39, 40-49, 50-59, 60-69, 70-79, 80-89, 90+. This grouping constitutes the finest common resolution of the above data sets.

Note that it makes a difference whether death counts are reported by Destatis on a weekly or daily basis. According to an ISO standard [26], a German statistical year consists of either 52 weeks (i.e. only 364 days taken into account) or 53 weeks (371 days), as is well reflected by the daily death counts (black spots in Figs. 4, 5) differing from weekly ones (orange squares in Figs. 4, 5). Accordingly, every five or six years, when a 371-days year is inserted in the official statistical data presentation, the weekly counts show a leap artefact: As compared to a regular calendar year containing 365 days (or 366 in a leap year), the missing one-day contributions (presently in Germany: about 2,800 fatalities) of the preceding statistical 364-day years are showing up due to summation over seven days more than in the years before.

### 2.3. Two models for calculating expected AMCs, and hence EMCs, for whole years and flu seasons

When looking at the time courses of age-cohort-specific weekly AMR values 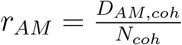 during the last twenty years (Fig. 1), we see that they were all tendentiously decreasing, bar rem stochastic fluctuations in the observed data, from the early 2000s until around 2014, from which on they seem to saturate to near constant values. Here, we propose two plain AMC models for estimating *expected* (‘regular’) AMC values in Germany, generally for an arbitrary time interval within a year. In our present analysis, we restricted ourselves to estimate expected *annual* AMC values (52 calendar weeks, i.e. 364 days) as well as *flu-seasonal* ones (33 calendar weeks from #40 to next #20 [25], i.e. 231 days). In fact, each expected-AMC model simply consists of the set of all cohorts’ AMR functions, which depend on just the observed (weekly if not daily) cohorts’ AMCs in the selected time interval within a year, the cohorts sizes *N*_*coh*_, and the time period chosen for fitting (here, the years 2000-2019). In the first, simpler model (termed ‘constant’), each cohort’s expected AMR value is a constant, the mean value over six years (2014-2019, see below). In the second, slightly more elaborate model (termed ‘exponential’), each cohort’s expected AMR was assumed to follow an exponential course with time. The function parameters are calculated by demanding this AMR function to best fit over the chosen period to the observed (time) course of AMR data points of which each is an arithmetic mean value across the selected interval within a year (here, either the year itself or a flu season). The AMR functions can be used (i) to interpolate (estimate) potentially missing AMR data points within the fitting period, (ii) to estimate expected AMR (and thus AMC) values at any data point within the chosen fitting period, or (iii) to extrapolate (prognosticate) expected AMR (and thus AMC) data points at times beyond 2019, the latter two being the model applications within this study.

**Figure 1:**
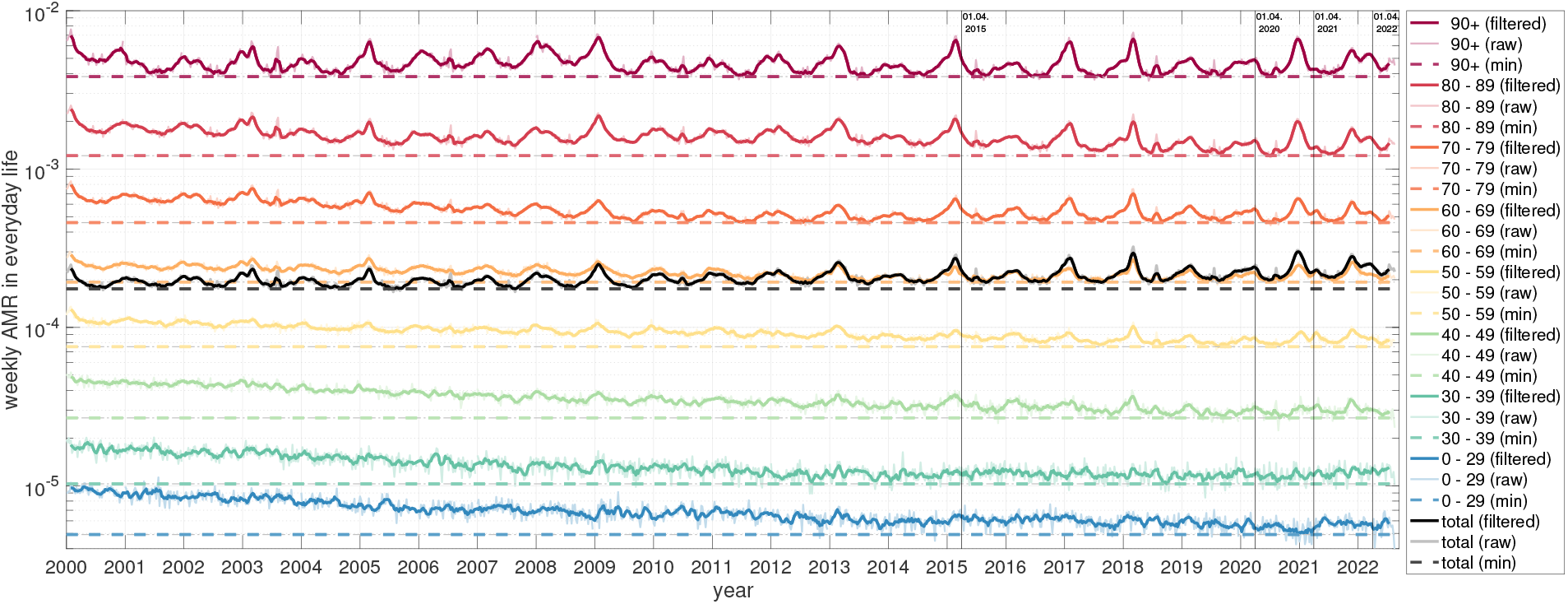
Weekly German AMR time courses between 2000 and 2022. Colours represent different age-cohort-specific as well as total (black) AMRs. Note that the raw data (dotted lines) had been smoothed (solid lines) here for better depiction by a moving average filter of a width of 5 weeks. The absolute minimum for each (filtered) weekly cohort AMR is indicated by a horizontal coloured dashed line in order to better observe temporal trends (see Fig. 4 and 5). Vertical solid lines are located at the first of April, i.e. the approximate end of the flu season, for 2015, 2020, 2021, and 2022.

In the simpler, constant model, we take advantage of the aforementioned saturation of the German AMR between 2014 and 2019 to calculate a mean annual AMR value for each cohort. We do so by fitting a constant function in a least-squares sense to the six data points, provided by the *fit* routine of the “Curve Fitting Toolbox” in MatLab (version R2021b). With this, we prognosticate the expected total German AMCs for the years 2020-2022 by calculating a weighted sum of the obtained (time-constant) AMR values, with each cohort’s size *N*_*coh*_ taken from the observed (time-dependent) demographic distribution in the prognosis year. Accordingly, in the constant model for expected AM, the course of the total population’s AMC over both the fitting and prognosis time periods is solely due to shifts of the demographic distribution with time (see Fig. 3).

In the more elaborate, exponential model, we take all available pre-pandemic years 2000-2019, again conducting a fitting procedure, to determine best fitting parameters (two for each cohort: *a*_*coh*_ and *b*_*coh*_) of the exponential AMR function

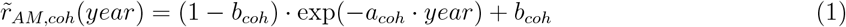

to the year-resolved *normalised* AMR data points of each age cohort separately, 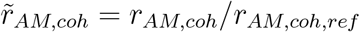. The normalisation parameters are the reference values *r*_*AM,coh,ref*_ = *r*_*AM,coh*_(2000), i.e. the cohorts’ AMR values observed in 2000. We then prognosticate the expected AMCs for the years 2020-2022 by extrapolating these exponential AMR fits and performing the same age-cohort AMR weighting in each prognosis year as for the constant model. For all of these annual estimates, we give 95% confidence intervals that account for the fluctuations in the observed AM data points, using the *predint* routine in MatLab.

Note that we generally summed 52 weekly observed AMC values in one year to calculate the annual AMC values, which serve as the input for fitting the AMR function parameters. Thus, the parameters of our expected AMC models represent exactly 52 weeks, i.e. a 364-day statistical year [26]. Therefore, any AMC model output is scaled by the factor 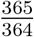 to calculate an AMC estimation of a regular year, and by 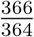 in a leap year.

The same model calculations can be easily applied to estimate expected AMC values of any time interval within a year. Here, we restricted ourselves to estimate expected AMC values for only one further, specifically selected interval within a year, namely, the ‘flu season’. Accordingly, the weekly (or daily if available) observed AMC values in a year have to be contracted by summation over only the interval of a flu season. We chose a fixed length of the latter by adopting its RKI definition [25, p. 13,17], namely, the interval from (inclusive) calendar week #40 in one year to (likewise inclusive) #20 in the subsequent one, and thus a flu season comprising 33 weeks, i.e. exactly 231 days. After contraction of the AMC values in each of the flu seasons 2000/2001-2019/2020 (short form: 2000/01-2019/20), the fitted exponential model parameter values (i.e. those of the cohorts’ AMR functions *r*_*AM,coh*_(*year*) from Eq. (1)), and thus the cohorts’ expected AMC data points of interest, are specific for flu seasons.

This method can be generally applied: A model for calculating, in a given population, expected AMC data points can be easily constructed for *any specific* time interval shorter than a year, not only a flu season. Within each cohort and at yearly resolution, just the cohort’s weekly (or daily if available) AMC values observed have to be summed over the chosen time interval. Subsequently, the fitting procedure provides parameter values (for either the cohort’s exponential function or simply a constant, the mean of the interval sums taken into account) that represent the (smoothed) course of the cohort’s expected AMC data points over the whole fitting period.

### 2.4. PFR values from registered SARS-CoV-2 “cases” and C19 death counts: Solving the problems of multiple tests per person and time delays

While displaying PFR values in 2020 and 2021, we noticed extremely low PFR from late 2021 on, as compared to the first-year SARS-CoV-2 infection period from May 2020 until summer 2021 (see Appendix A, Table A.7 for details). Our interpretation of these massively dropping PFR values is that, from some instant in late 2021 on, there has been an evidently inconsistent relation between official “case” counts (raw *PCR*_*pos*_, number of positive *tests*) and the number of positively tested *persons*. However, to calculate a meaningful PFR, the number of persons having died (*D*_*PF*_) must be divided by the number of *persons* tested positive (*PCR*_*pos*_ processed to represent *persons* rather than positive *tests*). In other words: Since late summer 2021, the officially published (raw) *PCR*_*pos*_ (and thus PFR) values can not be reasonably interpreted as representing positively tested persons without additional processing. Fortunately, such processing, which implies a re-interpretation of what *PCR*_*pos*_ counts mean, proved *possible*: We assume that the PFR values *until* summer 2021 are *disease characteristics*, which have been consistently determined due to a person-to-test ratio being nearly one-to-one in the first year of the ‘Corona pandemic’, i.e. from May 2021 to May 2022. With this, we re-calculated the raw *PCR*_*pos*_ values for the subsequent, second year of the ‘Corona pandemic’, i.e. May 2021 to May 2022, and its sub-period, the flu season 2021/22. We did so by multiplying the raw *PCR*_*pos,May*21*/*22_ values between May 2021 and May 2022 with the factor 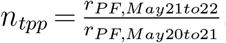, and accordingly the raw *PCR*_*pos,flu*21/22_ values for the flu season 2021/22 with 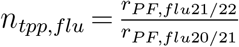. The results of this *PCR*_*pos*_ post-processing are presented in Sec. 3.3.

Another observation brought to light that reporting PFR values by dividing the number of C19 deaths per week (*D*_*PF*_) by PCR-positive “cases” per the *same* week (*PCR*_*pos*_) is not recommended for two reasons. First, the number of PCR tests can well differ greatly by the week, so there is significant scatter along week-resolved time courses. Second, from receiving the positive test result (or its date of registration) to the actually corresponding death event, there is usually a time delay of several days or even weeks [27]. Here, we show that taking raw PFR values from an unprocessed calculation of a same-week ratio overestimates, on average, the PFR values that arise when comparing death counts to the number of positive test results from days ago. Therefore, our PFR analysis started with finding realistic delay values between the courses of PCR-positive “case” counts and PFCs, one value for each age cohort. In doing so, we only relied on courses until mid of 2021, because later (raw) PCR-positive “case” counts turned out to be unreliable (see above). We determined these delay values in a straightforward way by systematical time shifting of the two data sets relative to each other, with covering the range from May 2020 to May 2021. To this end, in each cohort, (i) a (pseudo-)daily resolution was generated by applying cubic-spline interpolation to the courses of both the PFC values *D*_*PF*_ and the counts of PCR-positive “cases” *PCR*_*pos*_, all known as weekly sums; (ii) these two day-resolved courses were systematically shifted (in discrete day steps) relative to each other, and the arithmetic mean of the (daily) PFR values (the ratio 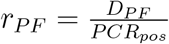 of counts in a time unit) was calculated for each day-shift value; (iii) the very delay value with the minimal mean *r*_*PF*_ value for a maximally five-week delay was identified (see Fig. A.7; mean *r*_*PF*_ values in Table A.7).

### 2.5. C19 death counts put into context: Analysing PFR(PFC) against the background of AMR(AMC)

As the final step within this study, we performed the analysis presented in Sec. 2.3 again cohort-wise and both annually, with yet the years adapted to the ‘Corona pandemic’ (i.e. across the calendar year boundaries: From May 2020 to May 2022), and for the two flu seasons in this two-year episode. To put PCR-conditional mortality (i.e. PFCs and PFRs) into AM context, we applied the exponential model estimation of the age cohorts’ AMRs (Sec. 2.3, Eq. (1)), while also knowing the *PCR-conditional* cohort sizes (for each, its *PCR*_*pos,coh*_ value), to the group of Germans that have received a positive PCR test, i.e. the whole PCR-conditional sub-group of the population. Within each cohort, we contrasted the number *D*_*PF,coh*_ of persons in the sub-sub-group therein, i.e. C19 fatalities, with the number *D*_*AM*|*PF,coh*_ of persons who were estimated by our AMC model (Eq. (1)) to have expectably died due to any cause within the PCR-conditional sub-group of *PCR*_*pos,coh*_ persons. That is, we calculated the PCR-conditional EMC by subtracting deaths *D*_*AM*|*PF,coh*_ expected according to our AMC model from the number *D*_*PF,coh*_ of official C19 deaths. PCR-conditional EMCs (here, annual or flu-seasonal values) quantify *excess* deaths in the sub-group of official C19 deaths, i.e. fatalities that are in excess of what is expected due to all causes. This method of determining a conditional EMC is generally applicable to *any* conditionally selected sub-group of a population. Within the sub-group of PCR-positive persons, we suggest to identify these definite excess deaths (PCR-conditional EMCs) with those having occurred *due to* CoViD-19. The results of this analysis are presented in Sec. 3.3.

## 3. Results

### 3.1. The AMR course over the last twenty years: A Simpson’s paradox

In this section, we will address the beneficial representation of AMR courses over time (see Sec. 2.3) and we show that the interplay of these courses with demographic changes result in a paradoxical phenomenon. Ignoring this interplay may result in devastatingly wrong forecasts of AMCs, as is discussed later.

In order to achieve our main goal, the valid estimation of EMCs, we start here with scrutinising age-cohort-resolved AMR time courses during the last twenty pre-pandemic years, with the age cohorts being determined by the available data sets (see Sec. 2.2). The first main finding of this investigation: The trend of the cohort’s AMR time courses may well be described by plain functions versus time (Sec. 2.3). In Germany since 2000, they turned out to be well of exponentially saturating character in all but one age cohort (Fig. 1; more obvious when normalised to each its 2000 value: Fig. 2b). The scientific value gained by identifying such basic functions for (model-based) AMC calculations lies in (i) their potential to reflect a trace of the dynamical mechanisms behind changing AM in a population, and (ii) the transparency they lend to AMR and thus AMC estimations. The latter benefit comes in a big part from the inherent character of any continuous function in time, particular if being of exponential character: to reflect physical and biological processes, rather than possibly arbitrary, mathematical black box assumptions. Due to time-wise continuity and moreover being low-parametric (determined by only two parameters for each twenty data points, see Appendix B for their values and confidence intervals), (iii) they also have inherently a smoothing, averaging character, which is a technical advantage for using such a function-(model-)based approach to estimate AMC *base lines*.

**Figure 2:**
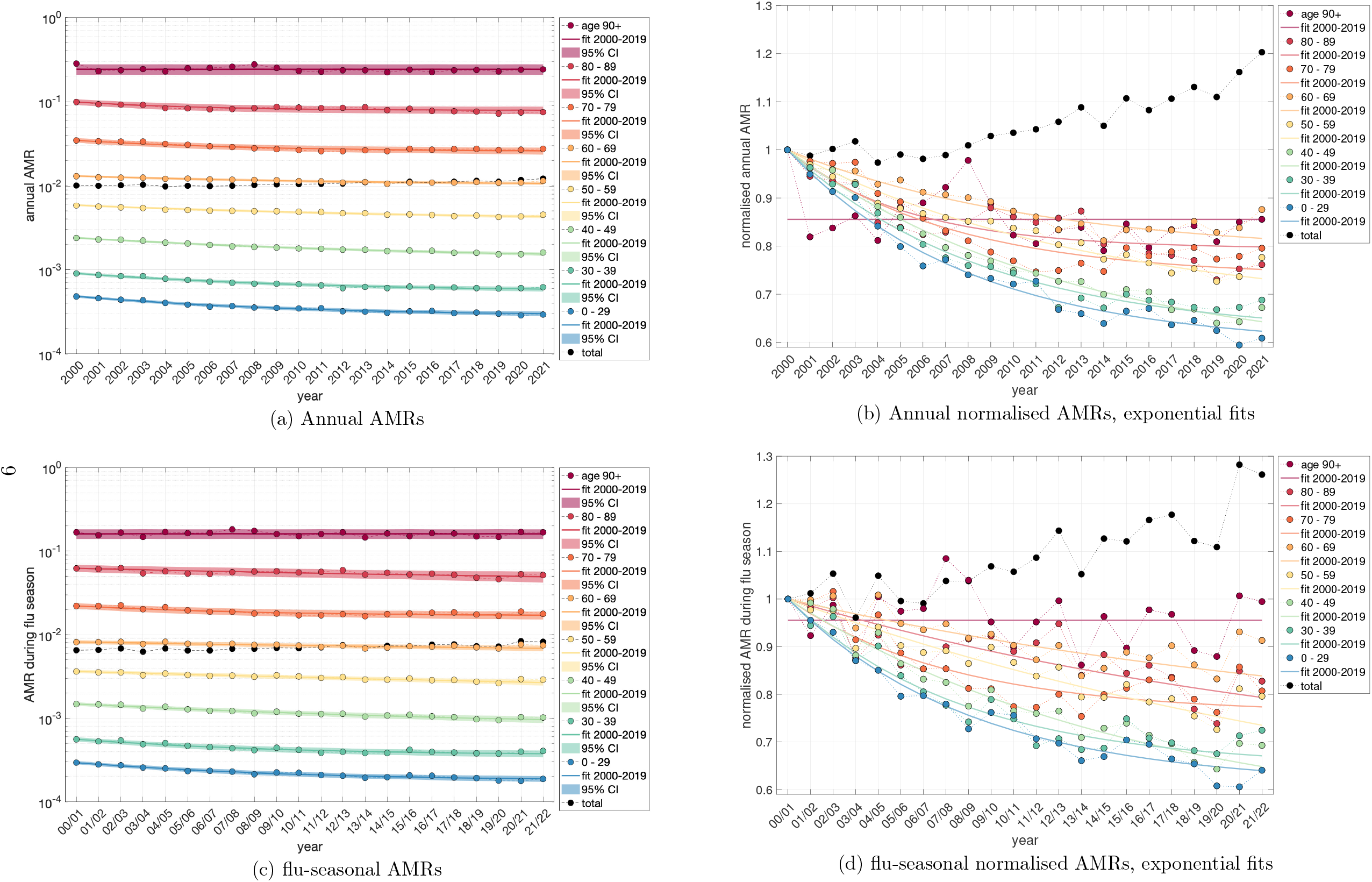
German annual (top) and flu-seasonal (bottom) AMR per age cohort from 2000-2021 and 2000/01-2021/22, respectively. The left panel contains the absolute AMR values together with exponential-fit confidence intervals; in the right panel, for a better depiction, all AMR values are normalised to the year 2000 (season 00/01) reference value, together with the exponential point estimate. See Appendix B for details on the parameter values.

The second main finding of this scrutiny: Resolving AMC to age cohorts matters. A year-wise contraction of each age cohort’s weekly AMR time course (Fig. 1) into a single data point, starting with the year 2000, provided a sequence of annual AMR values (coloured bullets in Fig. 2a). Their general trend with time can be better seen by plotting each cohort’s AMR course of the annual values normalised (Fig. 2b) to its reference value (the value in 2000 or in the 2000/2001 flu season Fig. 2d, respectively). Next, the course of the total German population’s AMR is calculated as the weighted sum over all cohorts’ normalised AMR values in a given year, with each cohort being weighted by its relative proportion (Fig. 3) in that year, times its respective reference value. If then both the year-resolved cohorts’ normalised AMR courses and the course of the total AMR (normalised to the sum of all cohorts’ sizes: the whole population size) are plotted over the last twenty years (Fig. 2b), a counter-intuitive phenomenon occurs in Germany: While each single cohort’s AMR tends to *decrease* over the years, in all age cohorts except the eldest whose AMR remains practically constant, the total AMR tends to *increase*, which applies consistently to both annual and flu-seasonal AMR courses.

**Figure 3:**
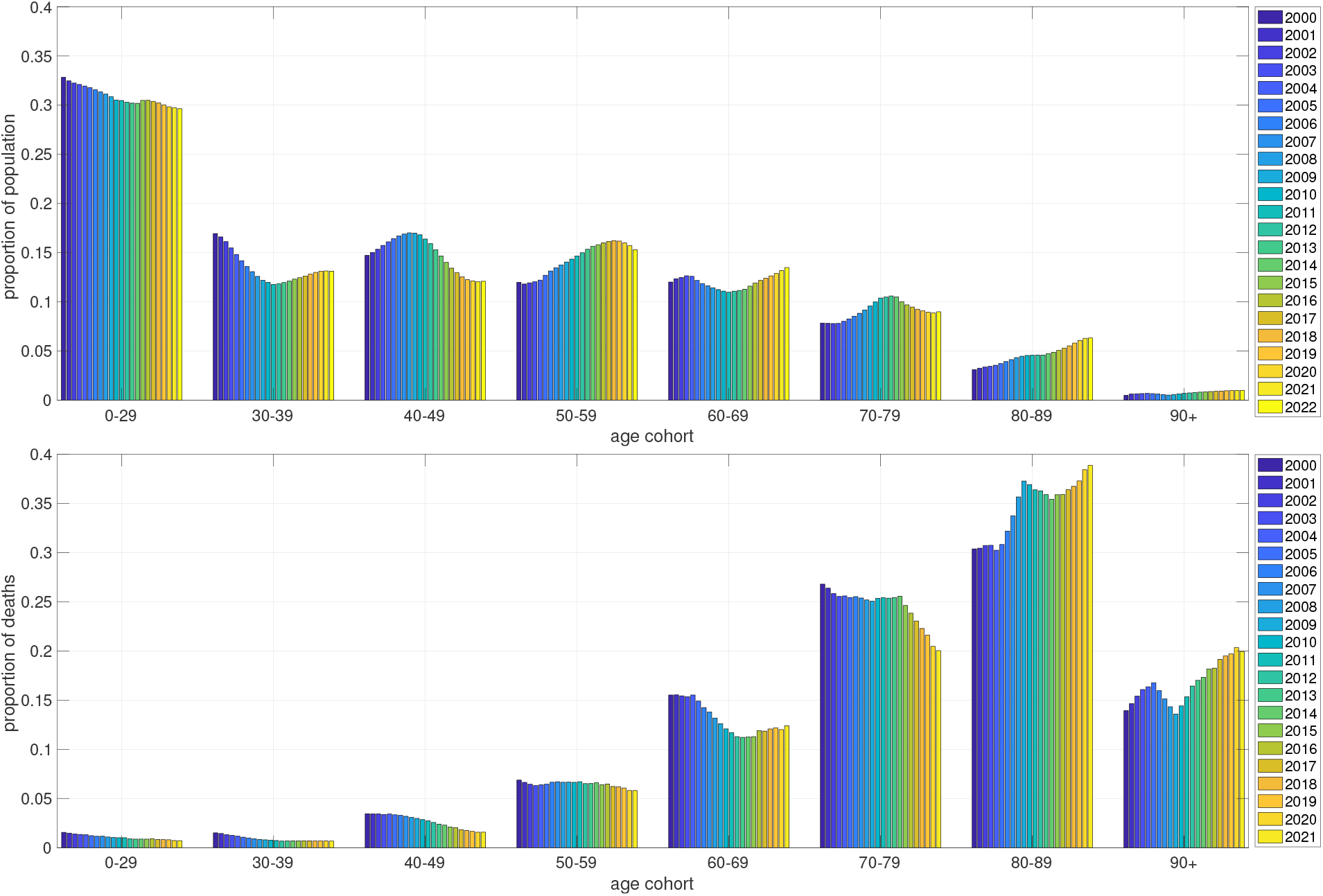
Demographic distribution of the German population’s relative proportions (in percent), between the years 2000 and 2022, of age cohorts (top panel) and death counts (bottom panel). In 2021, the officially stated number of German inhabitants was about *N*_*pop*_ = 83.5 million [32].

This phenomenon is known as *Simpson*’*s paradox* [28, 29, 30, 31]. It is explained by summing over sub-groups (cohorts) that differ in relative proportion (i.e. their weights, the time-dependent demographic distribution, Fig. 3) and effect strength (i.e. the AMR values, which are strongly age-dependent): Older people die at a higher probability per time (e.g. a year, Fig. 2a, or a flu season, Fig. 2c), and, in Germany during the last about ten years, the proportions of the 60-69, 80-90, and 90+ year old people increased, whereas those of all people younger than 60 years decreased (Fig. 3). Consequently, the (average) total probability of a German of whatever age to die within a year has increased, although each German below 90 years has gained more average life time during the last twenty years, until 2020 at least.

Note first that the exponential fits (according to Eq. (1)) in the 90+ cohort did not yield a meaningful result (no calculation of confidence intervals was possible), neither for the annual values in 2000-2019 nor for the corresponding flu-seasonal values; it was therefore substituted by simply fitting a *constant* for the 2000-2019 courses of each the annual and the flu-seasonal normalised AMR values (see Fig. 2b and Fig. 2d).

Note second, again, that only the data points between 2000 and 2019 (or flu seasons 2000/01-2019/20, respectively) have been used to calculate the fit parameters (separate for the years and the flu seasons). Thus, the line parts in Fig. 2b and Fig. 2d that pass the last two data points are extrapolations (prognoses) by the exponential fit functions (Eq. (1)). The overall graphs are the graphical representations of the cohorts’ *expected* AMR courses according to the exponential fit model, i.e. a smoothed (as compared to the fluctuating values observed), model-based (‘exponential fit’) reference for estimating EMCs. Not visualised here is the simplest reference model, the ‘constant fit’, solely consisting of horizontal fit lines of which each runs through the arithmetic mean of one cohort’s AMR values observed in the last six intervals (years or flu seasons) before 2020, i.e. a constant AMR value assigned to each cohort.

Note third that, according to Fig. 1, nothing else than ‘AM as usual’ occurred in Germany at any time since early 2020. Our assessment of entirely non-exceptional AMCs and all-cause EMCs in Germany during the SARS-CoV-2 era, be they assessed annually or during the flu seasons, is quantitatively substantiated in Sec. 3.2 (Tables 1, 2 in particular). Hence, it has to be assumed that any model calculation showing dramatic EMC in Germany during the last two years fell for the Simpson’s paradox and has to be carefully reconsidered regarding the shifts in the demographic distribution, i.e. changes of the age cohorts’ sizes, with time, see Sec. 4.2.

**Table 1:**
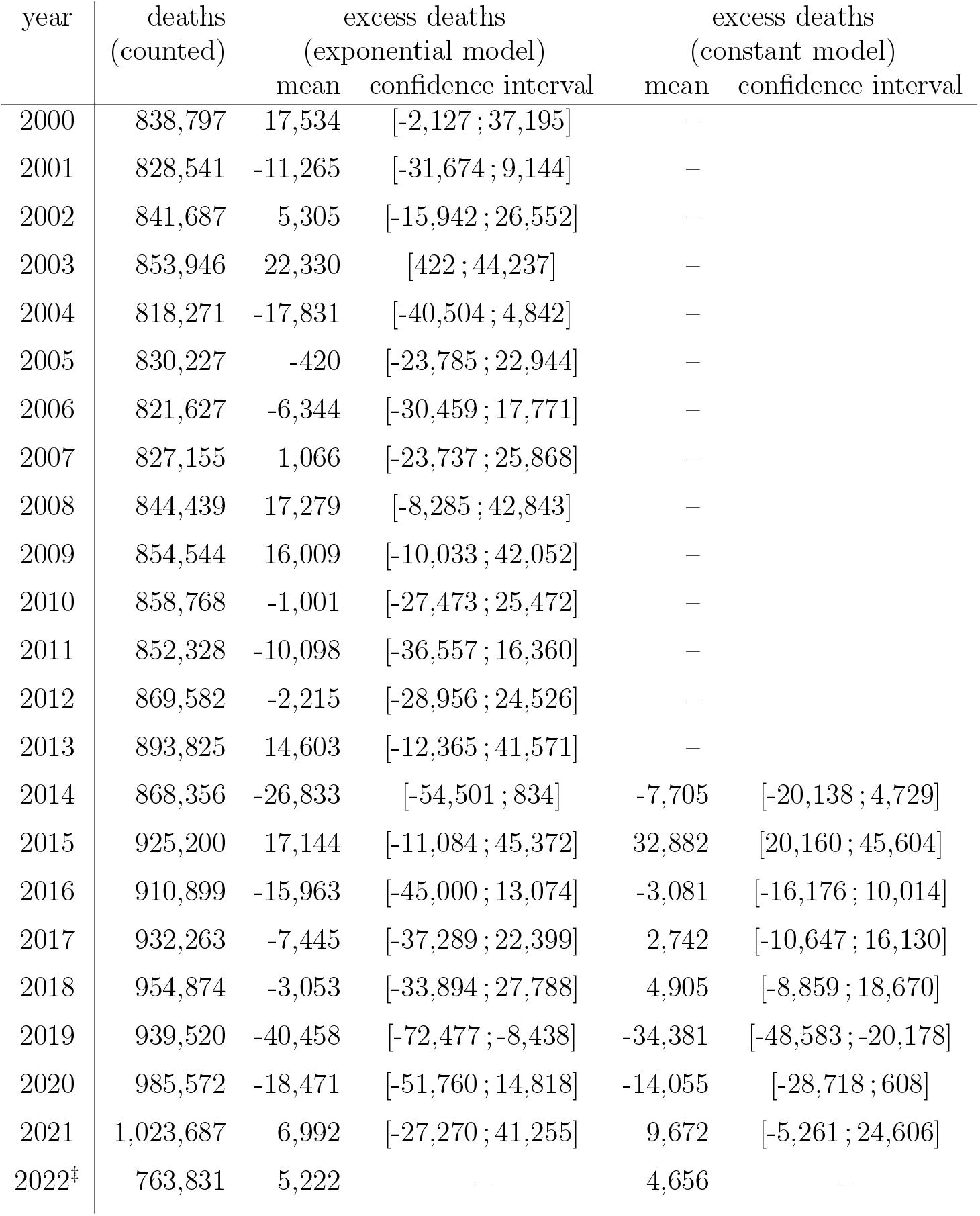
Counted AMC versus model-estimates (last three rows: extrapolation) in Germany during the years 2000-2021; ^‡^ until September 30th 2022 (273 out of 365 days), observed deaths officially registered plus 4,000 (estimate by empirical knowledge) due to be registered; population in 2021: *N*_*pop*_ = 83.5 million [32]; ‘excess’ (EMC) is counted minus model difference; for the constant model, under-mortality of -14,055 deaths in 2020 (reference: mean of 2014-2019 AMR values) correspond to -16.9 deaths per 100,000 persons, best comparable to Sweden with about -10 per 100,000 [33, fig. 2,bottom] (reference: mean of 2015-2019 AMR values); other than (short-term: 2015-2019) trend models applied to Sweden [33, fig. 2,bottom] (about +40 per 100,000), our (longer-term: 2000-2019) trend model (exponential) calculates slightly enhanced magnitude of prognosticated under-mortality: -22.1 per 100,000.

| year | deaths<br>(counted) | excess deaths<br>(exponential model) |  | excess deaths<br>(constant model) |  |
| --- | --- | --- | --- | --- | --- |
|  |  | mean | confidence interval | mean | confidence interval |
| 2000 | 838,797 | 17,534 | [-2,127 ; 37,195] | — |  |
| 2001 | 828,541 | -11,265 | [-31,674 ; 9,144] | — |  |
| 2002 | 841,687 | 5,305 | [-15,942 ; 26,552] | — |  |
| 2003 | 853,946 | 22,330 | [422 ; 44,237] | — |  |
| 2004 | 818,271 | -17,831 | [-40,504 ; 4,842] | — |  |
| 2005 | 830,227 | -420 | [-23,785 ; 22,944] | — |  |
| 2006 | 821,627 | -6,344 | [-30,459 ; 17,771] | — |  |
| 2007 | 827,155 | 1,066 | [-23,737 ; 25,868] | — |  |
| 2008 | 844,439 | 17,279 | [-8,285 ; 42,843] | — |  |
| 2009 | 854,544 | 16,009 | [-10,033 ; 42,052] | — |  |
| 2010 | 858,768 | -1,001 | [-27,473 ; 25,472] | — |  |
| 2011 | 852,328 | -10,098 | [-36,557 ; 16,360] | — |  |
| 2012 | 869,582 | -2,215 | [-28,956 ; 24,526] | — |  |
| 2013 | 893,825 | 14,603 | [-12,365 ; 41,571] | — |  |
| 2014 | 868,356 | -26,833 | [-54,501 ; 834] | -7,705 | [-20,138 ; 4,729] |
| 2015 | 925,200 | 17,144 | [-11,084 ; 45,372] | 32,882 | [20,160 ; 45,604] |
| 2016 | 910,899 | -15,963 | [-45,000 ; 13,074] | -3,081 | [-16,176 ; 10,014] |
| 2017 | 932,263 | -7,445 | [-37,289 ; 22,399] | 2,742 | [-10,647 ; 16,130] |
| 2018 | 954,874 | -3,053 | [-33,894 ; 27,788] | 4,905 | [-8,859 ; 18,670] |
| 2019 | 939,520 | -40,458 | [-72,477 ; -8,438] | -34,381 | [-48,583 ; -20,178] |
| 2020 | 985,572 | -18,471 | [-51,760 ; 14,818] | -14,055 | [-28,718 ; 608] |
| 2021 | 1,023,687 | 6,992 | [-27,270 ; 41,255] | 9,672 | [-5,261 ; 24,606] |
| 2022<sup>‡</sup> | 763,831 | 5,222 | — | 4,656 | — |

**Table 2:** Counted AMC versus model estimates (last two rows: extrapolation) in Germany during the flu seasons 2000/01 to 2021/22; population in 2021: *N*_*pop*_ = 83.5 million [32]; ‘excess’ (EMC) is count minus model difference; for comparison, estimated numbers published by the RKI [25, p. 47] are given in the last column.

| year | deaths<br>(counted) | excess deaths<br>(exponential model) |  | excess deaths<br>(constant model) |  | flu deaths<br>(RKI) |
| --- | --- | --- | --- | --- | --- | --- |
|  |  | mean | confidence interval | mean | confidence interval |  |
| 00/01 | 533,445 | 1,419 | [-20,065 ; 22,904] | — | — | — |
| 01/02 | 540,826 | -1,359 | [-24,029 ; 21,311] | — | — | 0 |
| 02/03 | 563,575 | 20,504 | [-2,874 ; 43,882] | — | — | 8,000 |
| 03/04 | 531,721 | -15,675 | [-39,898 ; 8,548] | — | — | 0 |
| 04/05 | 545,301 | -1,557 | [-26,327 ; 23,213] | — | — | 11,700 |
| 05/06 | 531,520 | -14,982 | [-40,173 ; 10,210] | — | — | 0 |
| 06/07 | 528,041 | -17,992 | [-43,499 ; 7,515] | — | — | 200 |
| 07/08 | 554,270 | 6,337 | [-19,522 ; 32,195] | — | — | 900 |
| 08/09 | 566,938 | 17,164 | [-9,068 ; 43,395] | — | — | 18,800 |
| 09/10 | 550,009 | -10,965 | [-38,095 ; 16,166] | — | — | 0 |
| 10/11 | 554,481 | -13,709 | [-41,526 ; 14,107] | — | — | 0 |
| 11/12 | 568,670 | -4,275 | [-32,581 ; 24,031] | — | — | 2,400 |
| 12/13 | 597,792 | 22,875 | [-5,771 ; 51,521] | — | — | 20,700 |
| 13/14 | 552,426 | -29,750 | [-59,068 ; -,433] | — | — | 0 |
| 14/15 | 614,852 | 24,600 | [-5,502 ; 54,702] | 33,913 | [21,110 ; 46,715] | 21,300 |
| 15/16 | 584,197 | -16,619 | [-47,727 ; 14,488] | -11,051 | [-24,249 ; 2,147] | 0 |
| 16/17 | 624,984 | 18,168 | [-13,826 ; 50,163] | 19,983 | [6,487 ; 33,479] | 22,900 |
| 17/18 | 632,810 | 16,980 | [-16,201 ; 50,162] | 15,080 | [1,213 ; 28,948] | 25,100 |
| 18/19 | 604,302 | -22,518 | [-57,178 ; 12,143] | -28,111 | [-42,404 ; -13,818] | — |
| 19/20 | 619,348 | -22,041 | [-58,579 ; 14,496] | -31,394 | [-46,198 ; -16,589] | — |
| 20/21 | 675,714 | 27,594 | [-10,547 ; 65,735] | 14,526 | [-,594 ; 29,646] | — |
| 21/22 | 683,195 | 30,443 | [-9,287 ; 70,173] | 13,692 | [-1,640 ; 29,025] | — |

### 3.2. Observed AMCs versus AMC-model-calculated fatalities: EMCs, annually and during flu seasons

In this section, we show that both the constant and the exponential model (Sec. 2.3) yield exceptionally good fits within the intervals 2014-2019 and 2000-2019, respectively, as well as realistic prognoses for AMCs in 2020 and 2021. Contrary to claims by the RKI of significant, and by the WHO even of drastic EM in Germany during the SARS-CoV-2 era we even find negative EMCs (‘under-mortality’).

In Fig. 4, we have plotted actually observed data points (counts on daily basis: black spots) and the correspondingly expected exponential model estimates (green triangles with 95% confidence intervals) of the total annual AMC since the year 2000. The annual sums of the daily observed AMCs (the only data available by day) are always within the 95% confidence interval of the exponential model fit, except for the year 2019 in which significantly fewer than expected people died. Accordingly, the exponential model generally well matches the observed AMCs over the whole analysed period from 2000 until today. For the flu-seasonal data in particular (Fig. 5), the AMCs expected from the simplest (constant) model (blue triangles with 95% confidence intervals) match the observed AMCs very well. All estimates by the constant model demonstrate impressively that, in Germany since 2014, the total AMC time course is dominated for the most part by shifts in the demographic distribution with time (Fig. 3). The differences between both models’ estimates, whether for whole years or flu seasons, are usually smaller than between an exponential model estimate and an observed AMC on a yearly basis. Note that the observed 2019/20 flu-seasonal AMC in Germany is significantly *below* both model estimates (Fig. 5), although this season contains the first SARS-CoV-2 wave of infection (start of the ‘Corona pandemic’).

**Figure 4:**
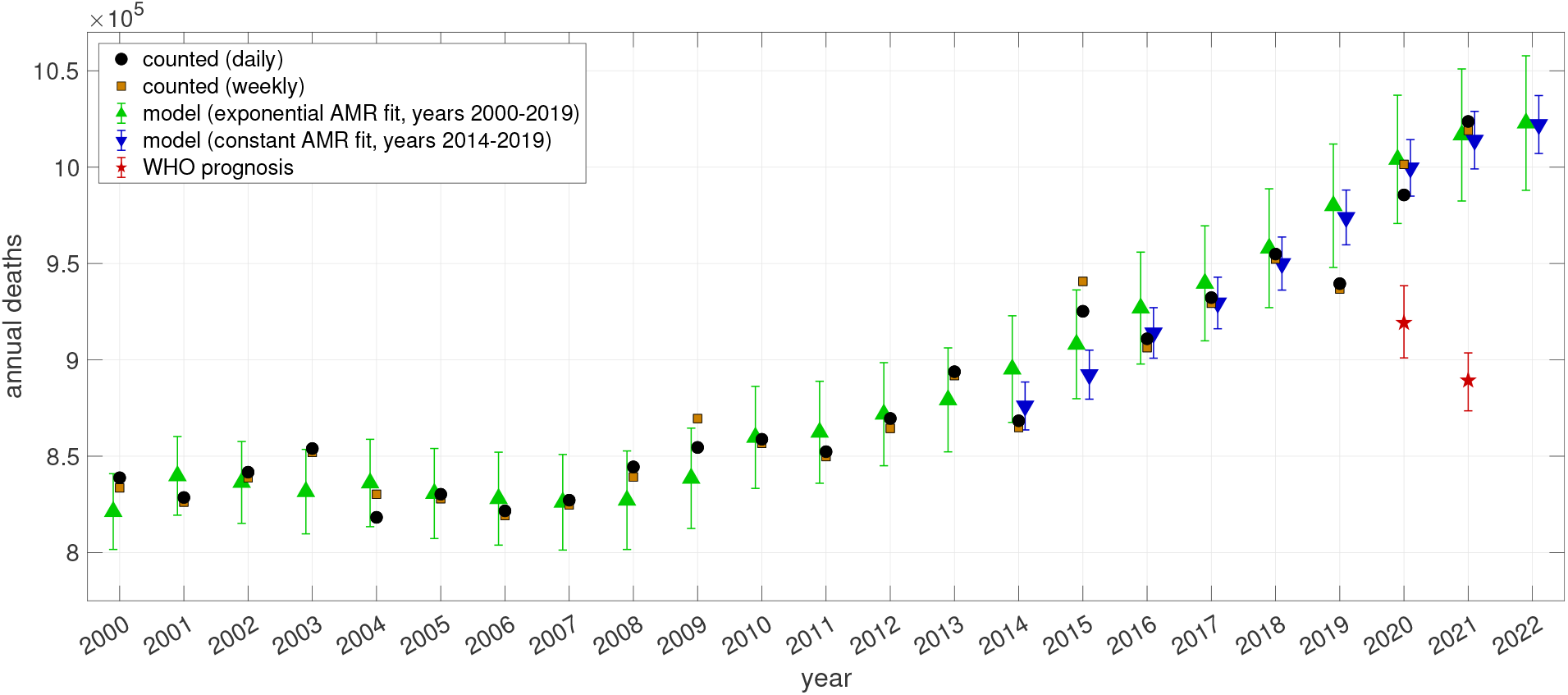
German data of counted annual deaths (derived from daily counts in black dots, from weekly counts in brown squares), versus model estimates or prognoses (exponential model in green upward-pointing triangles, constant model in downward-pointing blue triangles) including 95% confidence intervals for the years 2000-2022, versus WHO’s AMC prognosis for 2020 and 2021 ([15], red stars); population in 2021: *N*_*pop*_ = 83.5 million [32].

**Figure 5:**
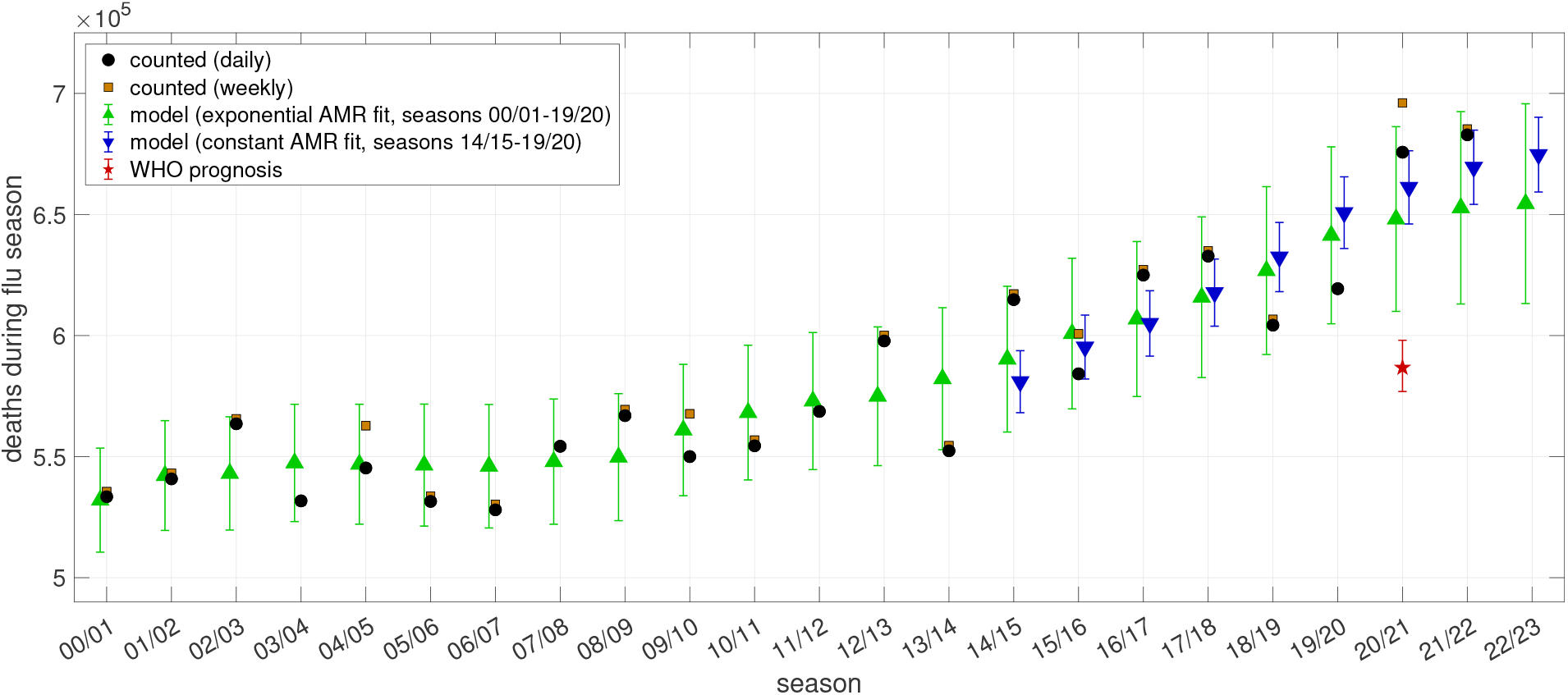
German data of counted flu-seasonal deaths (daily in black dots, weekly in brown squares), versus model estimates or prognoses (exponential model in green upward-pointing triangles, constant model in downward-pointing blue triangles) including 95% confidence intervals for the seasons 2000/01 until 2022/23, versus the WHO’s AMC prognosis for 2020/21 and 2021/22 ([15], red star)

For the total German population, we have listed in Table 1 the exact numbers (visualised in Fig. 4) of observed annual AMCs (‘deaths’) since 2000 and their corresponding differences (‘excess deaths’: EMCs) to both the exponential and constant model estimates. The observed AMCs are generally sums over 365 days, and 366 days in a leap year. The essential numbers are the EMC values in the last two rows of Tables 1, 2, and, by our analysis, we can make a clear statement: *Fewer* people than expected died in 2020 (negative excess value of -18,471), and in 2021, a mild EMC of about 7,000 showed up. In both ‘Corona pandemic years’, non-existing EM gives evidence of there having been none exceptional public health situation whatsoever. In contrast to our finding, the WHO prognosticated a total EMC of about 195,000 during 2020-2021 (red stars with 95% confidence intervals in Fig. 4). Note that the actual AMC data points for 2020 and 2021 are unfathomable 6.8 and 18.5, respectively, standard deviations away from the WHO’s calculated point estimates. The consequential non-validity of their model and the impact of missing age cohort distribution is discussed in Sec. 4.2.

In Table 2, the exact numbers (visualised in Fig. 5) of observed AMCs (‘deaths’) in Germany during the flu seasons—exactly 33 weeks, thirteen at a year’s end plus the first twenty of the next: 91 plus 140 days, or 141 in a leap year—are reported. The exponential (flu-seasonal AMR fit) model output representing 33 weeks (231 days) is scaled to a leap year by the factor 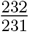, and, like in Table 1, the differences (‘excess deaths’: EMCs) to both the exponential and constant model estimates are also given. Like in the data for the calendar years, the essential numbers are the excess values in the last two rows. In the flu season 2020/21, an excess of 27,594 deaths can be attributed mainly to the second and third waves of putative SARS-CoV-2 infections. The excess of 30,443 deaths was even a little bit higher in the following season 2021/22. These excesses followed two seasons 2018/19 and 2019/20 with mortality below the expected value (negative excess values, i.e. ‘under-mortality’), at almost the same magnitude. This is evidently a typical pattern for seasonal flu waves: Sometimes up to four flu seasons of under-mortality in a row are followed by a similar sequence of stronger flu seasons with excess values on a level of 20,000 deaths. Since the last ten years, the magnitude of these fluctuations seem to slightly increase, with none of the flu seasons close to the expected value occurring in-between anymore. Looking back until 2000, the excess values in the two ‘Corona seasons’ have been just slightly higher than in the strongest influenza seasons of the last twenty years (2002/03, 2012/13, 2014/15). The two ‘Corona seasons’ were thus not at all on exceptional mortality levels. Our rating of flu seasonal excess mortality is reconfirmed by published RKI numbers [25, p. 47]: Since season 2000/01, they specified four excess estimates above 20,000 deaths, the highest (25,100) in the season 2017/18, which may be seen as a calibration value for an EMC occurring in a significant (moderately strong yet usual) flu season in the last decade. Notably, we found a higher excess than the RKI in the season 2002/03, and there are remarkable near-matches in 2008/09, 2012/13, and 2014/15 (referring to our exponential model at least). According to our analysis, the RKI excess values in 2016/17 and 2017/18 even seem to be slight exaggerations.

Interestingly, on the one hand, even under-mortality and a very low excess can be seen in the ‘Corona pandemic’ years 2020 and 2021, respectively. But, on the other hand, one can see significant excess deaths during the flu seasons 2020/21 and 2021/22. Correspondingly, there must have been significant under-mortality in the spring and summer periods in 2020 and 2021, which compensated flu-seasonal excess within few months, which we are investigating in the next section, regarding PCR-conditional deaths.

### 3.3. Putting PCR-conditional death counts into context: PFC against AMC background

This section addresses the question “Have people died with or due to SARS-CoV-2?”. For this, we simply put PCR-conditional death counts (PFCs) into the context of regularly expected, all-cause death counts (AMCs), i.e. present the results of calculating PCR-conditional EMCs (according to Sec. 2.5). They are based on the processing of raw PFC data (*PCR*_*pos*_ as observed) and corresponding PFR determination, as explained in Sec. 2.4. For each age cohort, we took its AMR values (Fig. 2), as well as its count of PCR-positive persons (*PCR*_*pos*_ processed) and its number of fatalities proclaimed to be related to SARS-CoV-2 (*D*_*PF*_) two weeks later, the latter two combined into PFR values (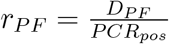; Fig. A.8 and Table A.7). As shown in Appendix A, the (raw) number *PCR*_*pos*_ of PCR-positive tests evidently has to be re-interpreted since mid of 2021 as compared to the start of the RKI publishing these data since early 2020: only until summer 2021, tests and persons are practically congruent, afterwards, *PCR*_*pos*_ must be processed to represent persons. All other data can be used as given above, the AMR values in particular, as they are only depending on plain, non-processed counts of all German citizens, and of all who died of any cause during the past: the plain registration of all deaths in files maintained by local authorities. Likewise, the PCR-conditional death counts *D*_*PF*_ seem to be consistent; in other words: The RKI criteria for proclaiming a death to be ‘related to SARS-CoV-2’ do not seem to have changed during the whole ‘Corona pandemic’ since early 2020. Such consistency can be most notably concluded from the fact that, after having applied our processing (see Sec. 2.4) to the officially published *PCR*_*pos*_ data since mid of 2021, the post-processed, inferred relative distribution of excess (PCR-conditional minus all-cause) death counts in 2021/2022 across age cohorts, given in Tables 5 and 6, are in very good proportion and accordance with those calculated without any *PCR*_*pos*_ processing in 2020/2021, given in Tables 3 and 4, respectively.

**Table 3:** Calculated German PCR-conditional (C19) EMCs (‘excess’) between May 2020 and May 2021; AMC model estimation: *D*_*AM*|*P F*_ ; SMR value: 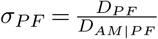.

| age cohort | $PCR_{pos}$ | $D_{PF}$ | $r_{PF}$ | $D_{AM\vert PF}$ | excess | $\sigma_{PF}$ |
| --- | --- | --- | --- | --- | --- | --- |
| 90+ | 69,573 | 17,526 | **0.2519** | 17,012 | 514 | 1.03 |
| 80-89 | 195,016 | 36,274 | **0.1860** | 15,582 | 20,692 | 2.33 |
| 70-79 | 180,201 | 16,588 | **0.09205** | 4,753 | 11,835 | 3.48 |
| 60-69 | 310,528 | 7,195 | **0.02317** | 3,367 | 3,828 | 2.14 |
| 50-59 | 552,075 | 2,616 | **4.738e-03** | 2,388 | 228 | 1.10 |
| 40-49 | 481,410 | 586 | **1.217e-03** | 749 | -163 | 0.78 |
| 30-39 | 518,435 | 186 | **3.588e-04** | 309 | -123 | 0.60 |
| 0-29 | 1,052,065 | 65 | **6.178e-05** | 320 | -255 | 0.20 |
| total | 3,359,303 | 81,036 | **0.02412** | 44,480 | 36,556 | 1.82 |

**Table 4:** Calculated German PCR-conditional (C19) EMCs (‘excess’) during flu season 2020/21; AMC model estimation: *D*_*AM*|*P F*_ ; SMR value: 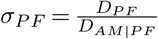.

| age cohort | $PCR_{pos}$ | $D_{PF}$ | $r_{PF}$ | $D_{AM\vert PF}$ | excess | $\sigma_{PF}$ |
| --- | --- | --- | --- | --- | --- | --- |
| 90+ | 68,608 | 17,328 | **0.2526** | 11,025 | 6,303 | 1.57 |
| 80-89 | 191,992 | 35,815 | **0.1865** | 9,474 | 26,341 | 3.78 |
| 70-79 | 176,979 | 16,306 | **0.09214** | 3,022 | 13,284 | 5.40 |
| 60-69 | 305,056 | 7,050 | **0.02311** | 2,092 | 4,958 | 3.70 |
| 50-59 | 540,003 | 2,550 | **4.722e-03** | 1,443 | 1,107 | 1.77 |
| 40-49 | 466,665 | 569 | **1.219e-03** | 447 | 122 | 1.27 |
| 30-39 | 501,518 | 180 | **3.589e-04** | 188 | -8 | 0.96 |
| 0-29 | 1,010,865 | 63 | **6.232e-05** | 190 | -127 | 0.33 |
| total | 3,261,686 | 79,861 | **0.02448** | 27,882 | 51,979 | 2.86 |

**Table 5:**
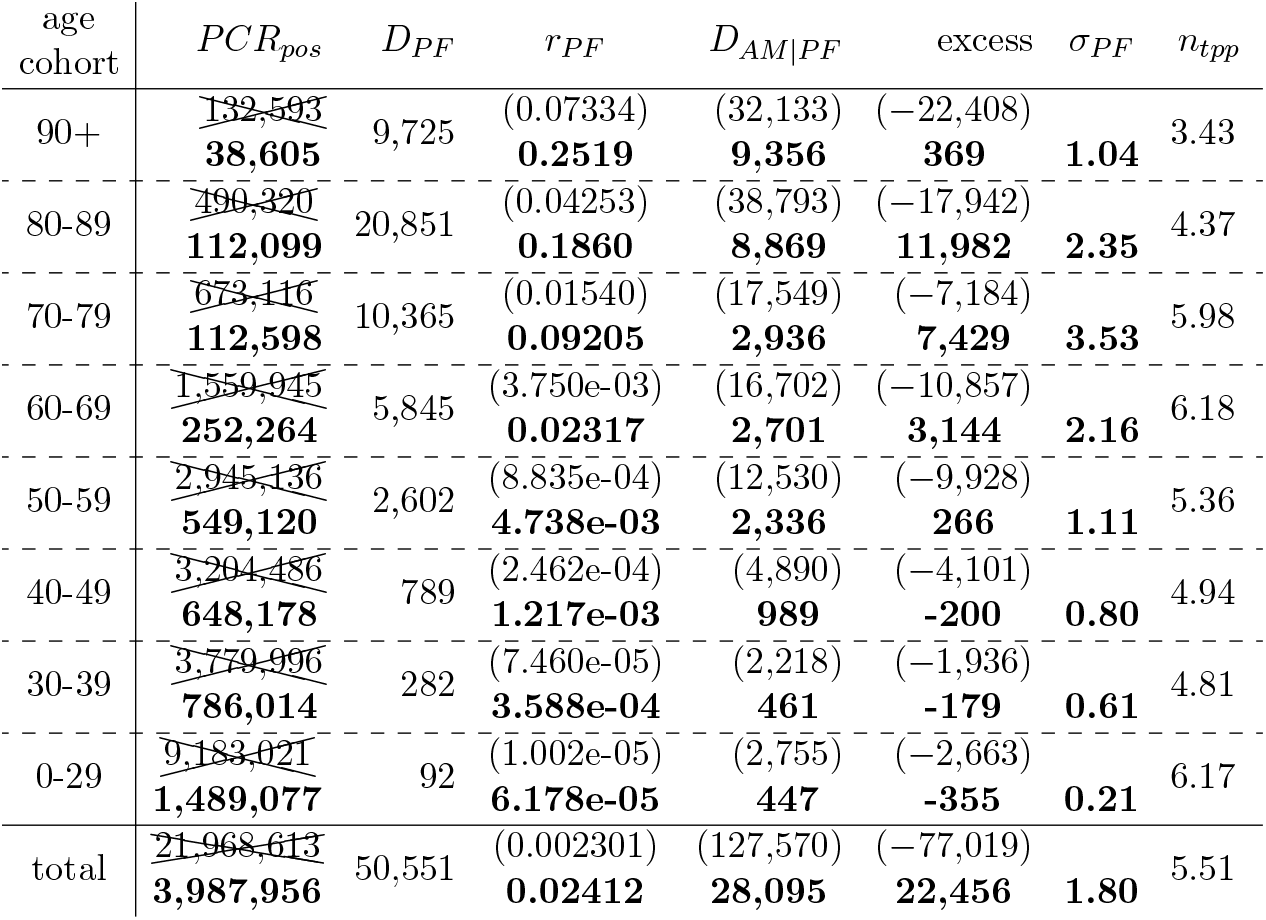
Calculated German PCR-conditional (C19) EMCs (‘excess’) between May 2021 and May 2022; multiple PCR tests: *n*_*tpp*_; AMC model estimation: *D*_*AM*|*P F*_ ; SMR value: 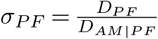. Bold numbers are due to *r*_*P F*_ being fixed to the value between May 2020 and May 2021 (Table 3).

**Table 6:**
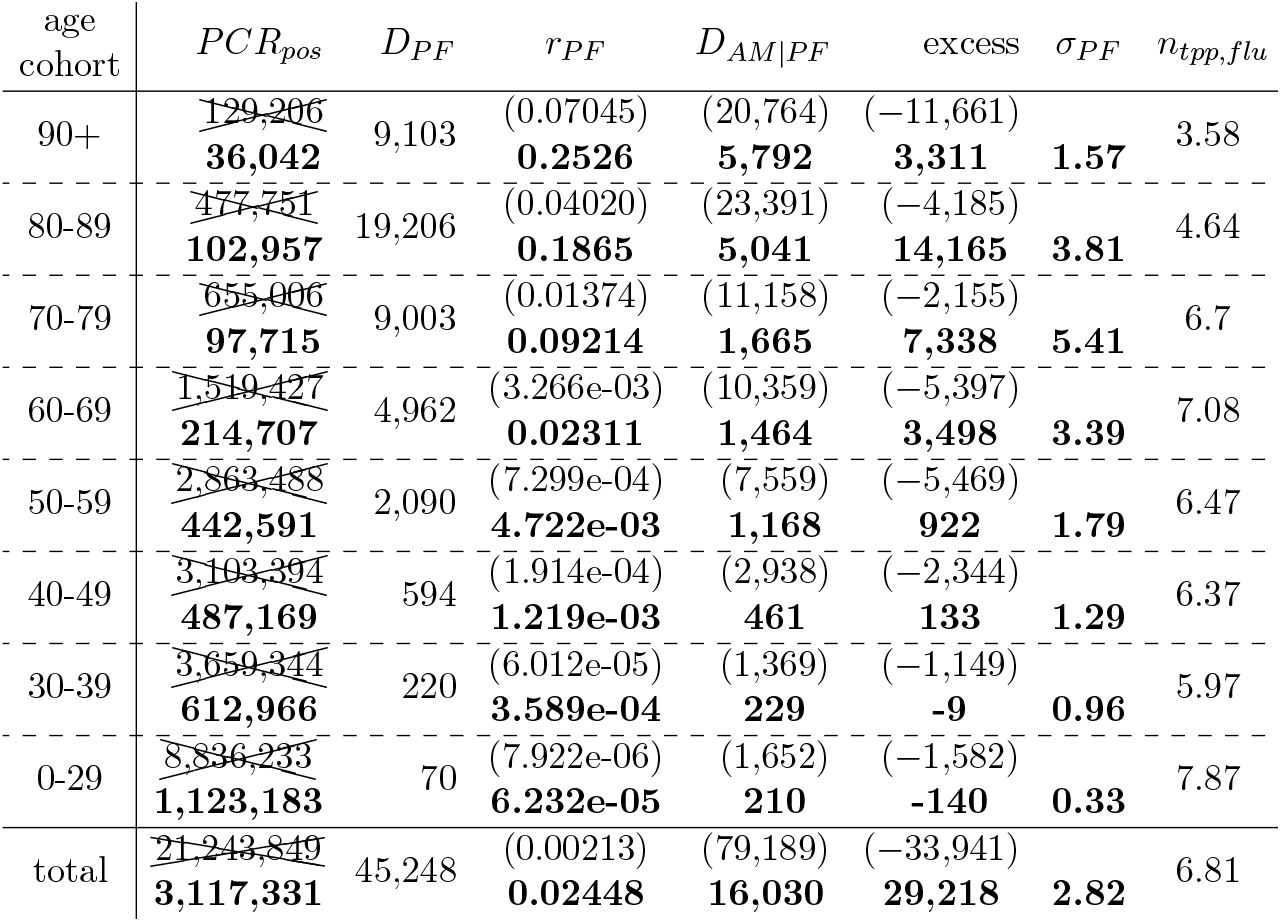
Calculated German PCR-conditional (C19) EMCs (‘excess’) during flu season 2021/22; multiple PCR tests: *n*_*tpp*_; AMC model estimation: *D*_*AM*|*P F*_ ; SMR value: 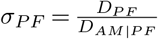. Bold numbers are due to *r*_*P F*_ being fixed to the value during the flu season 2020/21 (Table 4).

The PFR values for May 2020 until May 2021, practically identical to those in the corresponding flu season and taken as fixed disease characteristics, are printed in bold font in Tables 3 and 4. The result of the *PCR*_*pos*_ processing can be seen in Tables 5 and 6 in which we have crossed out the raw *PCR*_*pos*_ numbers, put all values depending thereon in round brackets, and printed all processed results also in bold font. The ratios *n*_*tpp*_ and *n*_*tpp,flu*_, which we would interpret as the number of multiple PCR tests per person within a year or flu season, respectively, are given in the last columns of Table 5 and Table 6, respectively.

When looking at the ‘Corona pandemic’, first-year period, May 2020 until May 2021, we can see by comparing Table 3 with Table 4 that there were practically vanishing infection dynamics from late May 2020 until end of September 2020, since all numbers in the whole-year period May 2020-May 2021 only minutely differ from the corresponding flu season. The main result is found in the last columns of Tables 3 and 4: For each cohort, the calculated deaths in excess (PCR-conditional EMC) of what is expected according to AMR. These EMC values are what *might eventually* be related to SARS-CoV-2 in an epidemiological sense. The total EMC value in the PCR-conditional sub-group in Germany from May 2020 until May 2021 was 36,556, which contrasts to the RKI-officially declared 81,036 ‘CoViD-19 deaths’. Adapting the RKI nomenclature of deaths ‘due to flu’ (“Todesfälle durch Influenza” [25, tab. 3], see also Table 2, last column) to the context of SARS-CoV-2, this total EMC value of 36,556 can be seen as the number of persons who have died ‘due to CoViD-19’, whereas one might say that 81,036 persons have died ‘with CoViD-19’. Notably, the total PCR-conditional EMC was clearly higher in the corresponding flu season 2020/21, namely, 51,979 deaths. This gives clear evidence that many of the persons who deceased in the flu season had only few months to live anyway, as an under-mortality must have occurred, even in the PCR-conditional sub-group of Germans, in both the summer periods 2020 and 2021. Further, from May 2021 until May 2022, the EMC value 22,456 was lower than the EMC from May 2020 until May 2021 (36,556, Table 5) and also lower than the EMC of the corresponding flu season 2021/22 (29,218, Table 6). That the PCR-conditional sub-group is definitely *not* representative of the whole German population, and therefore does not tell a lot about the total German infection dynamics in the ‘Corona pandemic’ first-year period, becomes evident from the total unconditional (i.e. all-cause) flu-seasonal EMC (27,594 with exponential model, or even just 14,526 with constant model, see Table 2) being clearly lower than the total flu-seasonal EMC in the PCR-conditional sub-group (51,979).

As a side note of having the raw counts of positive PCR tests post-processed in the intervals May 2021-May 2022 and flu season 2021/22, we can eventually interpret the resulting, processed *PCR*_*pos*_ values (bold numbers in Tables 5 and 6, respectively) once again as the numbers of positively tested persons; the corresponding numbers *n*_*tpp*_ and *n*_*tpp,flu*_ have then gained the meaning of the average number of PCR tests per person and year or flu season, respectively; their moderately cohort-dependent values are reported in the last columns of Tables 5 and 6. The annual *n*_*tpp*_ values range from 3.4 to 6.2, and *n*_*tpp,flu*_ from 3.6 to 7.9 in the flu season. It seems that the test frequency has been raised most notably in the flu season 2021/22.

We recapitulate: According to our model-based estimations, about 36,500 Germans might be seen as having died due to CoViD-19 from May 2020 until May 2021, encompassing the ‘second and third Corona waves’. In the interval from May 2021 until May 2022, another about 22,500 Germans might be seen as having died due to CoViD-19. The total annual PCR-conditional EMCs from May 2020 until May 2022 add up to exactly 59,012. Hence, in Germany, between March 2020 and mid May 2022, no more than about 64,700 persons may have died due to CoViD-19 (PCR-conditional EMC), including about 5,650 excess deaths during the ‘first Corona wave’ in March/April 2020. The latter number has been estimated from [34]: they calculated an all-cause EMC of 8,071, with their mean of 2016-2019 AM baselines being lower than ours averaged over 2014-2019; furthermore, the number (8,674) of ‘CoViD-19 deaths’ according to the RKI multiplied by 0.65 (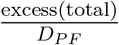 in Tables 4, 6) is about 5,650. Another view on Fig. 1 supports these statements: the strong influenza seasons in 2002/03, 2012/13, and 2014/15 show comparable patterns to the ‘Corona waves’.

The advantage of analysing the period between May 2020 and May 2022, instead of the whole years 2020 and 2021 as in Sec. 3.2, is that the former covers solely the ‘Corona pandemic’ situation. Again applying the RKI nomenclature, sums in the last rows of Tables 3 and 5 indicate that 59,012 persons died ‘due to CoViD-19’. As a side note, this differs by only about 2,000 from the PCR-conditional EMC of 56,911 for the roughly five-months-earlier period of the same length, covering exactly 2020 and 2021. Further resolving Tables 3 and 5 into age groups reveals that these deaths are almost exclusively attributable to the age groups 60+. Within the age group of 50-59 years, less than 500 fatalities can be ascribed to CoViD-19 and for age groups below 50, the negative excess numbers would be logged as overall zero deaths ‘due to CoViD-19’. In this light, the 255 deaths ‘due to CoViD-19’ during the flu seasons in the age group 40-49 (Tables 4 and 6), can be interpreted as people who must have had pre-conditions to die anyway within the next half of a year.

## 4. Discussion

### 4.1. Synopsis of our estimates of all-cause and PCR-conditional EMCs in the light of otherwise reported values

We suggest that our *PCR-conditional* EMC, which is calculated from RKI and Destatis data only, represents the very number of persons who have died *due to* CoViD-19. For Germany, we estimate that 56,911 persons have died due to CoViD-19 during the two complete two-years period 2020 and 2021. This total PCR-conditional EMC accounts for roughly 50% of the 114,866 deaths officially counted by the RKI during this two-years period as *related to* CoViD-19 [22] (2020: 44,039, 2021: 70,827; weeks #1-34 in 2022: 30,259). However, we found even a *negative* total *all-cause* EMC of -11,479 (resting solely upon Destatis data: Table 1) in those two years; a minor EMC of 6,992 in 2021 and moderate under-mortality of -18,471 in 2020. The correspondingly estimated all-cause EMCs for the flu seasons (Table 2) 2020/21 and 2021/22 have shown values of 27,594 and 30,443, respectively, which can be characterised as two typical, moderately severe flu seasons having occurred back to back. They followed however two essentially skipped flu seasons 2018/19 und 2019/20, with EMCs of -22,518 and -22,041, respectively. The comparison of annual with flu-seasonal AMCs implies that deaths occurring during these flu season were most likely to be due within the next months anyway. This view is strengthened by the observation of practically all of those deceased due to CoViD-19 being older than 60 years (Tables 3-6), accepting that all-cause EMCs were dominated by PCR-conditional EMCs (51,979 and 29,218, respectively), and the PCR-conditionally selected sub-group reflecting a sub-population of lower health on average. Further note that the all-cause EMC values (Tables 1, 2) calculated by our simplest, constant AMC model are even less ‘dramatic’ than by the exponential one, when referring to the flu seasons in particular. The high consistency of both model estimates strengthens the evidence of exactly non-exceptional ‘Corona pandemic’ mortality in Germany. Exceptionally deadly population dynamics in accordance with the WHO’s before-2009 description of what should be classified as an ongoing pandemic (in 2003-2009 comprising the qualifier “… resulting in epidemics world-wide with enormous numbers of deaths and illness.”) [35, 36, 37] are not discernible in Figs. 4, 5 and Tables 1, 2. Yet, it would seem common sense that only an “exceptionally severe disease” [38] spreading world-wide could be a decent reason for declaring a pandemic additionally labelled as “public health emergency of international concern” [39].

The official WHO reports announced a (*calculated*) mean of 66,794 excess deaths for 2020, and 128,194 for 2021 [15, 16, 17]. This notable, arguably pandemic-worthy EMC sum of 194,988, as well as the corresponding confidence intervals, however, lie far away from our estimates and the real numbers (Figs. 4, 5), and this sum was eventually reduced to 122,000 in an updated preprint [18, p. 21, and suppl. 6.7, p. S37, fig. S42]. However, this latter number, now being comparable to the 114,866 RKI counts, was neither resolved for years, nor were deaths ‘due to’ distinguished from ‘with’ CoViD-19, nor has the public data set [15] been updated accordingly. We found another two articles reporting EMCs for Germany between 2020 and 2021, which differed by a full order of magnitude; first, a two-paper study from German and Swedish scientists [40, 41] who proposed a total excess of 6, 317 + 23, 399 = 29, 716; second, an article by the “COVID-19 Excess Mortality Collaborators” [42], funded by the “Bill & Melinda Gates Foundation”, who claimed the highest EMC, namely, a mean 203,000 within a narrow range from 193,000 to 210,000.

What follows is a critical review of these vastly different EMC predictions and their underlying modelling procedures.

### 4.2. Shortfalls and consequences for modelling reliable EMC

At the end of the previous section, we have found that four mathematical models found four inherently different all-cause EMCs for Germany between 2020 and 2021: from -11,500 (our study) to 30,000 [40, 41] to 122,000 ([18], reduced from the official WHO report of 195,000 [15]), to 203,000 [42]. In this section, we aim at resolving how these different estimates arose from a prima facie unambiguous data set of a German AMC. Note that these EMCs are independent of any PCR-conditional counts and should thus not be equated or confused with deaths ‘due to CoViD-19’ (cf. last paragraph of Sec. 3.3) –at least, a priori. Three crucial model constituents are juxtaposed:

1. The data basis, including the range of years considered
2. Model equations and (over-)parametrisation
3. Age cohort distribution and demographic changes

*Ad 1)* We compare the underlying data sets of all four models. As stated in Sec. 2.2, we used German AMC data of 2000-2019 (exponential model) and 2014-2019 (constant model), respectively, as an input taken from Destatis [14, 21]. The Wang et al. model [42] likewise took the same Destatis data, but of 2016-2019 only [14]. De Nicola et al. [40] used Destatis data as well, but life tables (annual AMRs instead of AMCs) of 2017-2019 [43]. The Knutson et al. model [18] as an exception relied on AMCs of 2015-2019 taken from the Human Mortality Database [44]. While the different data sets should in principle comprise the same (or at least comparable) AMC data, all models were based on different time intervals. A look at Fig. 5 suggests that this choice may well determine the reliability of model outputs. Whereas 2014 was a year of rather low AM, 2015 was above the AM expectation due to our exponential model, 2016-2018 showed a little below-average mortality, and 2019 was a year of significant under-mortality. Now, the less years before 2020 are considered in any kind of fitting, smoothing, or superposing, the higher the impact of the outlier 2019, i.e. the trend to generally under-estimate AM-model-based *expectations* and thus report over-estimated EMCs. In the extreme case of choosing the interval 2015-2019 and taking weekly counts (Fig. 4), a mere linear fit of annual AMCs might well lead to even a slightly negative overall trend of prognosticated AMCs, if not considering the demographic changes (see below). Tracing the effect on our constant model, we predicted in Table 1 an under-mortality in 2020 and 2021 combined of -4,383 persons when using a six-year AMR average (2014-2019). If we switched to four-year (2016-2019) or three-year (2017-2019), respectively, averaging then we would end up with EMCs of 11,976 or 15,268, respectively.

*Ad 2)* We sketch the underlying models. The Knutson et al model [18] (as well as the initial WHO model [16]) and the Wang et al. model [42] employed a Poisson sampling approach to calculate estimated AMCs for 2020 and 2021 on the basis of their respective data sets. Both approaches combined the observed AM fluctuations around the sampled mean with spline-based tends to take account of annual as well as seasonal AM changes. While the Wang et al. model [42] used linear splines for seasonal changes and cubic splines for yearly (“secular”) trends, the WHO model in its initial form [16] used cubic splines for seasonal changes and thin-plate splines for yearly trends. Due to evidently the German and Swedish excess estimates in [16] having initially been “too high” [18, pp. 21, 37], the cubic-spline part was changed in the Knutson et al model [18] to hence employing linear trends for yearly changes in Germany as well. What is more in the Wang et al. model [42], the placement of the last knot for their splines was varied in order to yield four different predictions. With the implementation of another two sub-models, a Poisson regression model part and a “last-year” (2019) model part, the Wang et al. model finally consists of a weighted mean of six sub-models. In the De Nicola et al. model [40], life tables (AMR) are directly converted into expected AMCs, while using three differently detailed approaches (see next paragraph). Thereby, the years 2017-2019 contributed equally to forming the prediction for 2020, for both yearly and weekly data, with only applying the most elaborated of the three approaches. For an immediate comparison, we recall our model approach to calculate an expected AMC: we estimate age-cohort-specific AMRs of 2020 and 2021 by extrapolating exponential fits to the AMR courses between 2000-2019 (or, alternatively, constant fits between 2014-2019) multiply these extrapolated AMRs each by its proportion of the demographic distribution as predicted by Destatis [24], and sum all so-weighted cohorts’ AMRs.

By looking at the required parameters, we find that the De Nicola et al. model required no additional parameters, but rather assumptions on how life tables were to convert to future predictions of AMC. In our two model alternatives, we required either two exponential or one (constant) fit parameters per each of the eight age cohorts, together with each cohort’s AMR reference value in 2000 as a non-fit parameter, in order to fit the exponential or constant functions. We underline that both formulations arose from a careful look at the normalised AMR courses over the last 20 years (Fig. 2b) and may not be applicable to the situation in any other country. Yet, other plain, elementary, or special functions may well be suitable. For the initial WHO model [16] the Knutson et al. model [18] or the Wang et el. model [42], we find no definite number of required parameters. However, as cubic splines require at least as many as four parameters more than there are control (data) points, plus parameters for the sampling, plus additional sub-models in the Wang et al. model, the suspicion of quite some *over-fitting* arises. While the Poisson sampling method has the general advantage of being flexibly utilisable in cases of data transfer from countries being incomplete, the predictions for future developments have to be taken with caution. Wang et al. correctly state [42, suppl.] that cubic-spline extrapolation is subject to large uncertainties. This may have been the reason for the WHO model to eventually use linear splines [18], although a concrete reason their assessment “excess estimate [being] too high due to a combination of data/model issues” [18, pp. 21, 37] remains unresolved. In any case, the low observed AMC in 2019 seems to have driven the drastic under-estimation of 2020 and 2021 AMCs and hence drastic over-estimations of EMC (see the first model constituent above).

*Ad 3)* Now turning to the third crucial model constituent, we showed in Sec.3.1 that age cohort distribution and demographic changes play a crucial role in the prediction of future AMCs. Evidently, a Simpson’s paradox yields an increasing average AMR since 2000, while the trend is decreasing within each age cohort. Our AMC model as well as the De Nicola et al. model [40] included the demographic changes in Germany, the latter additionally including age cohort transition within years (Lexis diagram). The initial WHO model [16] and likewise the Knutson et al. model [18] do not include age cohort resolution, but a posteriori distributions of deaths within the cohorts can be calculated by them. The Wang et al. model [42] does not contain any age cohort or demographic considerations at all.

Summarising, our model produced the least deviations between modelled and measured AMCs, or, in other words, the least absolute EMCs. This had been achieved by (i) using a suitable long data history, (ii) assessing the Germany-specific time course of all age cohorts’ AMRs, (iii) using a low-parametric and straightforward model, and (iv) accounting for time-dependent age cohort distribution and thus demographic effects. In former times, the RKI had introduced similar model calculations [45], also applied to estimate flu-seasonal EMCs [46], and latest to the 2017/18 season [25, p. 13,17]; but up to date any EMC estimation by the RKI during the ‘Corona pandemic’ period is owing. We urge German authorities to not rely on generic world-wide prediction models, but provide a transparent and comprehensible estimate of EMCs of 2020 and 2021 as has been done for prior flu seasons, cf. Sec. 3.3.

The urgent need for a reliable estimate of, first of all, *all-cause* EMCs on basis of all-cause AMCs holds particularly as all of the above WHO-related reports [16, 42, 18] relate EMC to solely ‘CoViD-19’. However, no distinction in deaths ‘due to CoViD-19’ and ‘with CoViD-19’ in the light of PCR-conditional mortality (cf. Sec. 3.3) has been made down to the present day (of this article published). The herein presented methodology may provide a blueprint for other countries and serve as a benchmark in Germany for both reliable EMC estimates and a distinction between concomitant and causative ‘C19 deaths’.

## 5. Conclusion

World-wide media coverage of the ‘CoViD-19 pandemic’ is unsurpassed by any event recently (or probably ever). Accordingly, a plethora of data have been analysed to yield information, statistics, and key performance indicators of various kind. The dramatic number of increased mortality was one of the most important arguments by politicians to impose strict socio-economical measures against the population. In this work, we aimed for scrutinising the reportedly high excess mortality during 2020 and 2021. In stark contrast to the WHO estimate of 195,000 excess deaths in Germany in those two years, we even found a net under-mortality of -11,500, which implies that nothing but ‘dying as usual’ has happened, at least, when looking at the net proceedings across all age cohorts. Our analysis showed that the WHO estimates are flawed in terms of missing age-cohort-specific all-cause mortality characteristics, which obscure an underlying Simpson’s paradox. Further, all WHO-estimated excess deaths are barely subtly attributed to solely ‘CoViD-19’, be it by the afore-cited authors of WHO-endorsed modelling studies, WHO representatives, or German authorities and representatives of civil organisations and institutions. We have suggested a clear criterion to distinguish, at least on the epidemiological level, between deaths caused ‘due to CoViD-19’ or were just accompanied ‘with CoViD-19’. Based on thorough analysis of PCR-conditional mortality, out of the almost 115,000 ‘C19 deaths’ proclaimed by the RKI for 2020 and 2021 together, only about 57,000 or roughly 50% may be considered causatively related to CoViD-19.

Several open questions arise from our analysis, requiring thorough subsequent investigation: Do estimates of excess mortality for other countries suffer as well from ignoring demographic developments? How could such obvious miscalculations under the auspices of the WHO become accepted on a large scale? Shouldn’t political measures undergo reassessment in light of realistic and reliable excess mortality counts now being available? Why have none calculations of the kind presented here been conducted by German authorities as of the end of 2022, at least?

## Data Availability

All data produced are available online as indicated in the text.

## APPENDIX

### Appendix A. The course of the PFR during the SARS-CoV-2 era: Delay, tests, and persons

The results presented in this section are based on the calculations introduced in Sec. 2.4. In particular, we calculated a general time delay for all age cohorts from the time of tested infection to (possible) death. Figure A.6 shows the time courses of the age cohorts’ (raw) weekly PFR values 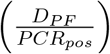 during the period of (putatively only) SARS-CoV-2 infections, as calculated by dividing the number of C19 deaths *D*_*PF*_ per week by the number of positive PCR tests *PCR*_*pos*_ per the same week. Next, Fig. A.7 shows the mean of PFR values 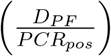 per age cohort when shifting the course of the registered death counts *D*_*PF*_ by up to 5 weeks forward in time with respect to course of the registered counts of positive tests *PCR*_*pos*_. For each age cohort, we see that the PFR attains a minimum between 4- and 24-day shifts (leaving away those younger than 20 years). For both simplicity as well as clarity reasons and because, on the one hand, almost all minima were very flat, and, on the other hand, another study [27] calculated a C19-”case”-death delay of 13 days in Germany for those older than 60 years, around which our values of all older than 50 years scatter (Fig. A.7), we generally chose a 2-week delay, which results in the weekly PFR values displayed in Fig. A.8.

**Figure A.6:**
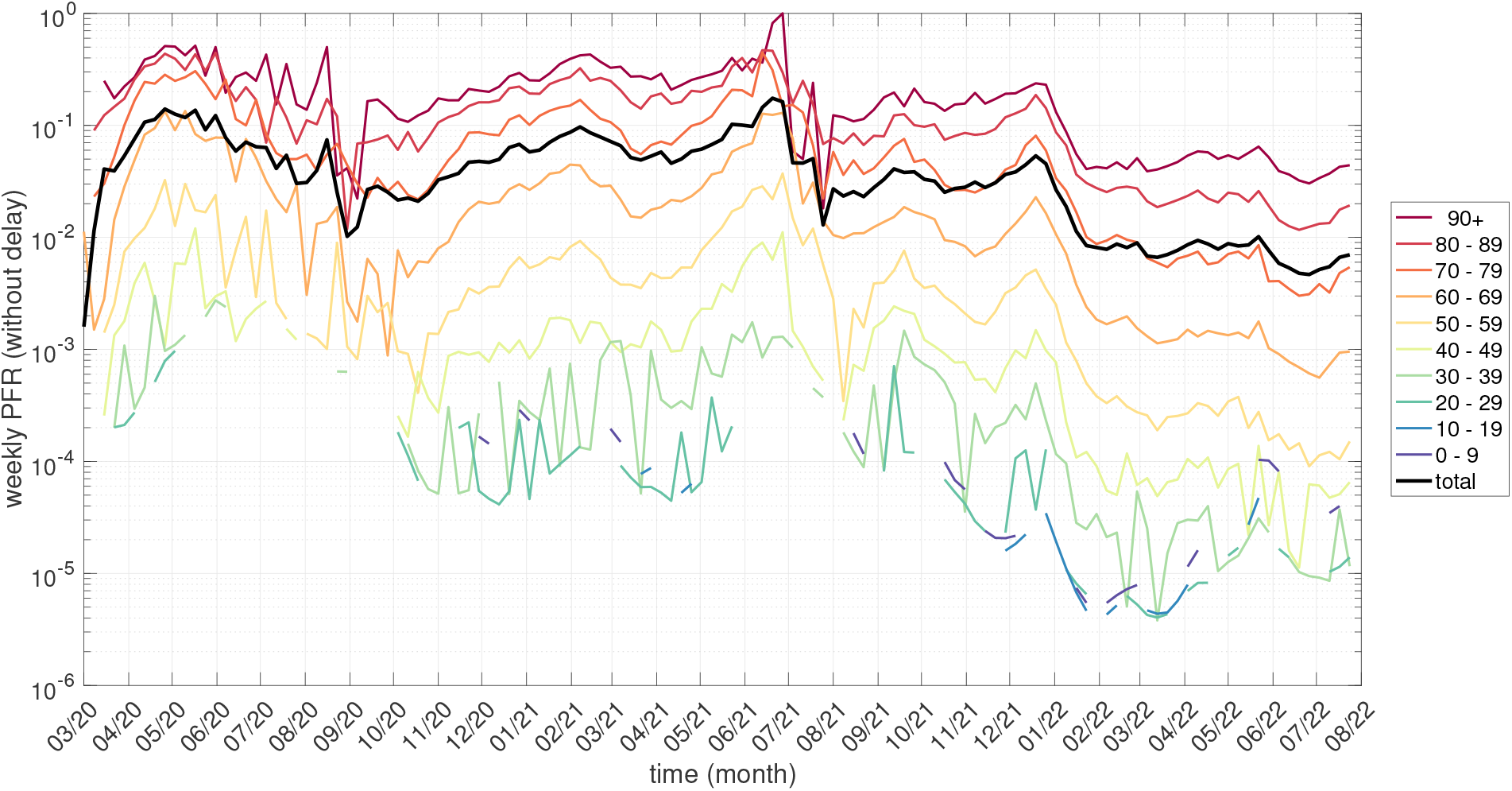
For each German age cohort, the raw, week-resolved PFR values in the period 05/2020 till 07/2022, i.e. weekly ratios 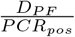 of the PCR-conditional (CoViD-19) death counts *D*_*P F*_ to the number *PCR*_*pos*_ of PCR-positive tests, as published by German authorities.

Two things strike to the eye in Fig. A.8. First, there were events of sudden PFR dipping by about an order of magnitude in early October 2020 in the two cohorts of those older than 80 years, preceded by each a peak in mid September. This is presumably an artefact simply due to very low numbers of both fatalities and PCR-positive persons. Second, after a similar peak-dip event between the end of July and late August 2021, the PFR values generally started to drop, first just slightly since October, and then the drop becoming particularly evident since the start of December 2021.

The effect of the PFR values in late 2021, even more in 2022, and in the flu season 2021/22 being so clearly lower than in 2020 and the season 2020/21, respectively, becomes evident in Table A.7 in which arithmetic mean values (annual and seasonal) of the fluctuating time courses in Fig. A.8 are given. Our suggestion to interpret such generally and significantly dropping PFR values is that there has been a near one-to-one relation between “case” counts (*PCR*_*pos*_) and positively tested persons *only until* summer or maybe autumn 2021. Afterwards, multiple PCR testings per person and year must have become a rule. We picked up on this issue in Sec. 3.3 in which we used the interpretation of multiple testings to implement an additional *PCR*_*pos*_ data treatment that infers processed PFR values for 2021 and 2021/22, which prove absolutely reasonable.

**Figure A.7:**
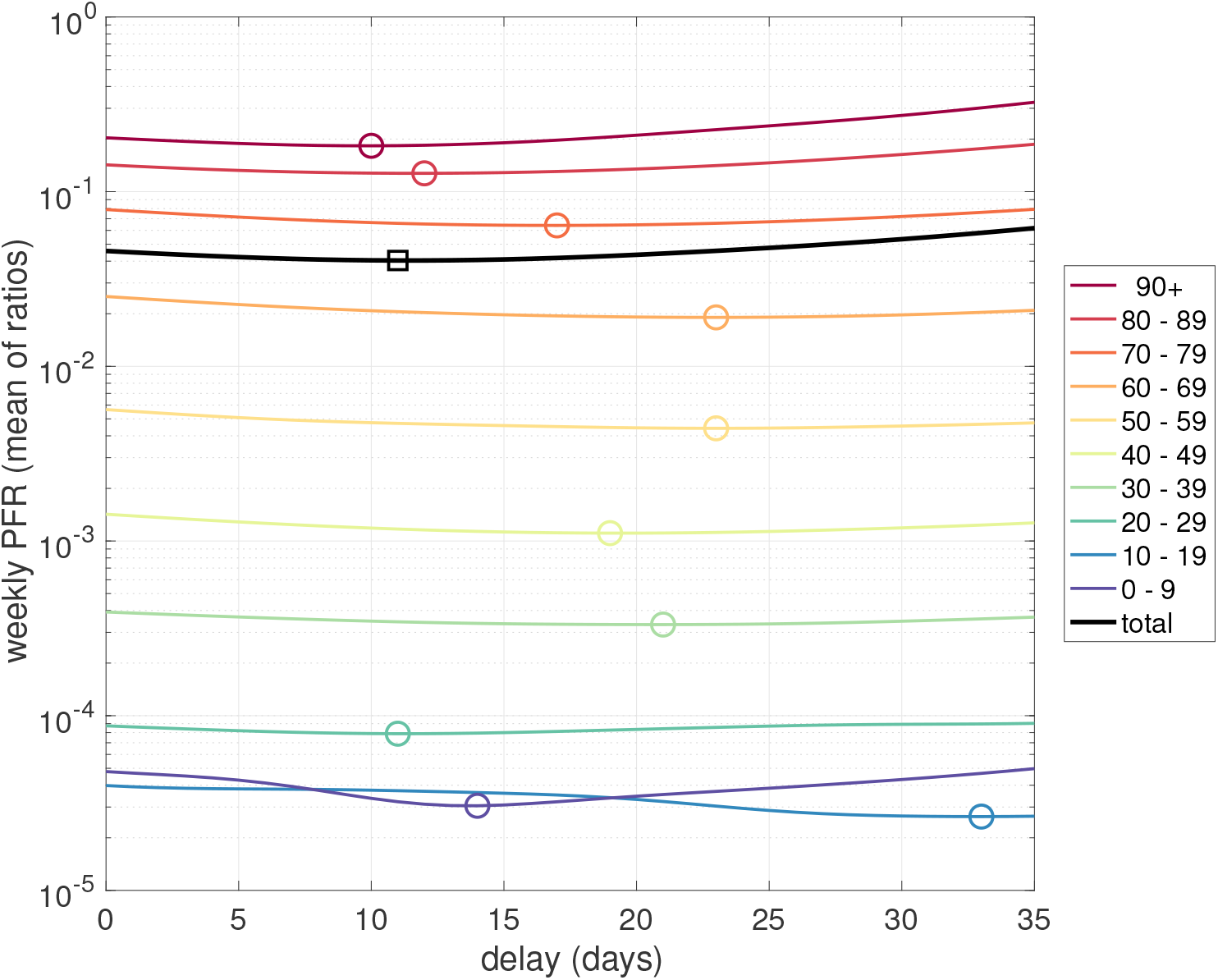
For each German age cohort, the mean of its daily PFR values (averaged across the period 05/2020 till 05/2021: Sum of daily ratios 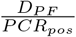 divided by the number of days taken into account) versus a varied time shift of the *D*_*P F*_ course forward with respect to the *PCR*_*pos*_ course; both courses registered in week resolution were interpolated through cubic splines for emulating day resolution; minima are indicated as circles or a square.

In Table A.7, we have juxtaposed PFR and AMR values (annual and flu-seasonal), *r*_*PF*_ and *r*_*AM*_, respectively, for the years 2019-2021 and the flu seasons from 2019/20 to 2021/22, respectively. The AMR values of the flu seasons were scaled by 52/33 (33 weeks per flu season), symbolised by 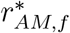, to make them comparable to both the annual AMR and the PFR values. Interesting observations in Table A.7 are summarised in the following; the first three items are about AM; the subsequent ones compare AM 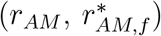 and PCR-conditional death rates (*r*_*PF*_), that is, the likelihoods of a German citizen to die within a year (or flu season scaled to a year, respectively) due to any cause (unconditional: *r*_*AM*_ or 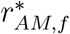, respectively) and to die within the respective interval (year or flu season, respectively) after having been tested positively in a PCR test (conditional: PFR). Due to the reasons given in the preceding paragraph, PFR values of 2021 and the flu season 2021/22 are not addressed here.

- All AMR values slightly increasing from 2019 to 2021 are fully consistent with the correspondingly increasing total EMC values evident from Table 1. The AMR values in 2020 and 2021 were higher than in 2019 for those with the highest AMR values (90+ and 80-89 cohorts), only slightly higher down to the age of 30, and even minutely lower for the 0-29 cohort.
- The 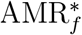 values of the flu (‘second and third Corona waves’) season 2020/21 are, in all cohorts 60+, moderately higher than in the flu season 2019/2020, slightly higher for the 30-59 cohort, and even minutely lower for the 0-29 year old. The very same holds for the classification of the flu season 2020/21 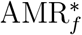 when compared to the annual AMR values both in 2020 and in 2021. This, most notably, owes to the fact that the flu season 2019/20 was exceptionally mild (see again the total excess values in Table 2), with its 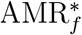 values being in any cohort even lower than the annual AMR values in all 2019-2021, except for the extraordinary low 2019 AMR in the 80+ cohorts: Not only was the flu season 2019/20 very mild, but also the preceding year 2019 was obviously extraordinarily mild (see again Table 1 indicating a marked total under-mortality, negative EMC, in 2019).
- The 2021/22 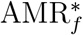 values are practically the same as in the season 2020/21, across all cohorts, which is again fully consistent with the respective EMC values being almost the same in these seasons, according to Table 2.
- In 2020, the PFR value of the oldest (90+: 0.2927) is higher than all AMR and 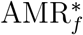 values in 2019-2021 and 2019/20-2021/2022, respectively. Interestingly, the PFR value in the flu season 2020/21 was lower than in the year 2020, and even lower than the respective 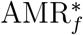 values in both flu seasons 2020/21 and 2021/22.
- The PFR values of the three cohorts 60-89 were all between two and three and a half times higher than the respective AMR values in 2019 and 2020, and 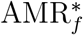 values in the flu season 2020/21, respectively. Again interestingly, like in the 90+ cohort, the PFR values of these three cohorts were lower in the flu season 2020/21 than in the year 2020.
- For the 50-59 cohort, the PFR values in 2020 and the flu season 2020/21 were just minutely higher than the respective AMR and 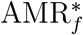 values. For all younger cohorts, the PFR values in both intervals were clearly lower (only a fourth among the youngest) than all AMR values from 2019 to 2021 and the 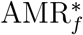 values of the flu seasons 2020/21 and 2021/22.
- The *total* (weighted by cohort proportion) PFR values in Germany were roughly double as high as total German AMR values, as they were dominated by the PCR-conditional death rate excess (PFR *>* AMR) of the four 60+ cohorts in 2020, and the three 60-89 cohorts in the flu season 2020/2021 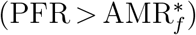.

**Figure A.8:**
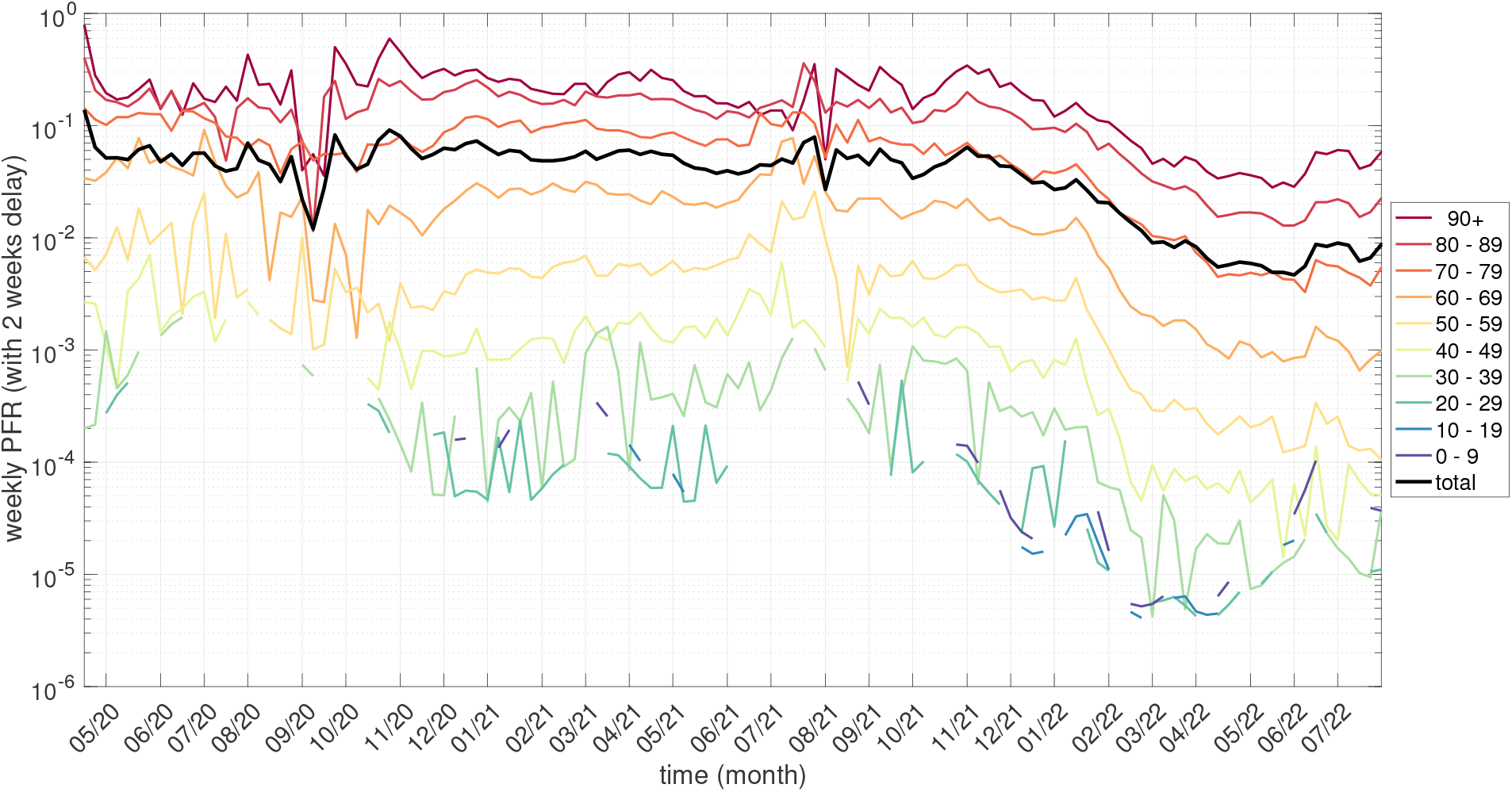
For each German age cohort, the week-resolved PFR values 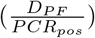 in the period 05/2020 till 07/2022, generally applying a forward time shift of two weeks to *D*_*PF*_.

**Table A.7:**
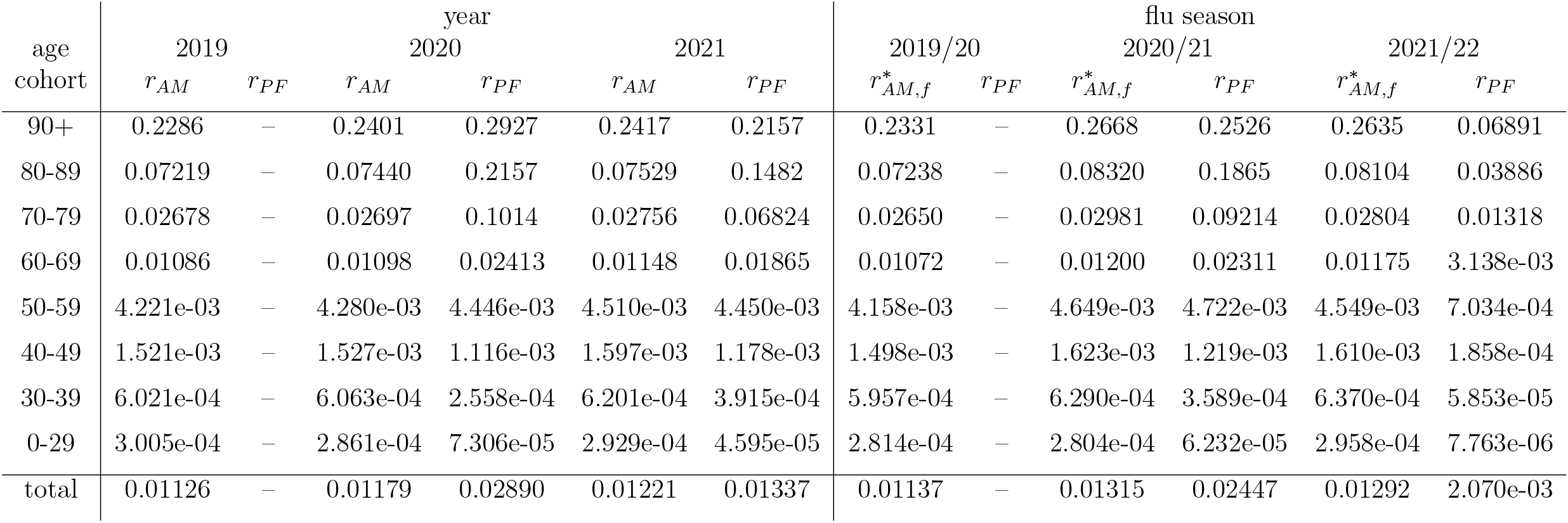
Calculated mean German AMR and PFR values (*r*_*AM*_ and *r*_*PF*_, respectively) from 2019 to 2021, including the flu seasons 2019/20, 2020/21, and 2021/22.

### Appendix B. Optimal fit parameters

**Table B.8:** Annual AMRs plus confidence intervals, calculated by both the constant and the exponential model, see Sec. 2.3. For the exponential model, function parameters are given as well. ^†^ For the age cohort 90+, a constant instead of an exponential fit was conducted

| age<br>cohort | constant model<br>AMR 2014-2021 |  | exponential model |  |  |  |
| --- | --- | --- | --- | --- | --- | --- |
| | mean | confidence interval | function parameters<br>$r_{AM,coh}(2000)$ | $\begin{pmatrix} a_{coh} \\ b_{coh} \end{pmatrix}$ | mean | confidence intervals |
| 0-29 | 3.113e-04 | [3.075e-04 ; 3.151e-04] | 4.813e-04 | $\begin{pmatrix} 0.1237 \\ 0.5927 \end{pmatrix}$ | 2.998e-04<br>2.981e-04 | [2.908e-04 ; 3.088e-04]<br>[2.890e-04 ; 3.073e-04] |
| 30-39 | 6.135e-04 | [6.085e-04 ; 6.184e-04] | 9.015e-04 | $\begin{pmatrix} 0.1148 \\ 0.6162 \end{pmatrix}$ | 5.865e-04<br>5.831e-04 | [5.689e-04 ; 6.041e-04]<br>[5.652e-04 ; 6.011e-04] |
| 40-49 | 1.620e-03 | [1.591e-03 ; 1.650e-03] | 2.376e-03 | $\begin{pmatrix} 0.07947 \\ 0.5599 \end{pmatrix}$ | 1.528e-03<br>1.513e-03 | [1.492e-03 ; 1.563e-03]<br>[1.476e-03 ; 1.549e-03] |
| 50-59 | 4.399e-03 | [4.346e-03 ; 4.452e-03] | 5.809e-03 | $\begin{pmatrix} 0.07299 \\ 0.6587 \end{pmatrix}$ | 4.255e-03<br>4.224e-03 | [4.164e-03 ; 4.345e-03]<br>[4.131e-03 ; 4.318e-03] |
| 60-69 | 0.0109 | [0.01083 ; 0.01098] | 0.0131 | $\begin{pmatrix} 0.08945 \\ 0.7829 \end{pmatrix}$ | 0.01069<br>0.01065 | [0.01047 ; 0.01091]<br>[0.01043 ; 0.01088] |
| 70-79 | 0.02702 | [0.02673 ; 0.02732] | 0.03464 | $\begin{pmatrix} 0.1369 \\ 0.7365 \end{pmatrix}$ | 0.02603<br>0.02596 | [0.02482 ; 0.02723]<br>[0.02474 ; 0.02718] |
| 80-89 | 0.07753 | [0.07598 ; 0.07908] | 0.09886 | $\begin{pmatrix} 0.1881 \\ 0.7944 \end{pmatrix}$ | 0.07893<br>0.07886 | [0.07496 ; 0.08289]<br>[0.07487 ; 0.08285] |
| 90+ | 0.2316 | [0.2285 ; 0.2347] | 0.2825 | 0.8555<sup>†</sup> | 0.2417 | [0.2248 ; 0.2585] |

**Table B.9:** Flu seasonal AMRs plus confidence intervals, calculated by both the constant and the exponential model, see Sec. 2.3. For the exponential model, function parameters are given as well. ^†^ For the age cohort 90+, a constant instead of an exponential fit was conducted.

| age<br>cohort | constant model<br>AMR 2014/15-2021/22 |  | exponential model |  |  |  |
| --- | --- | --- | --- | --- | --- | --- |
| | mean | confidence interval | function parameters<br>$r_{AM,coh}(00/01)$ | $\begin{pmatrix} a_{coh} \\ b_{coh} \end{pmatrix}$ | mean | confidence intervals |
| 0-29 | 1.956e-04 | [1.911e-04 ; 2.002e-04] | 2.939e-04 | $\begin{pmatrix} 0.123 \\ 0.6102 \end{pmatrix}$ | 1.880e-04<br>1.870e-04 | [1.808e-04 ; 1.951e-04]<br>[1.797e-04 ; 1.942e-04] |
| 30-39 | 3.918e-04 | [3.851e-04 ; 3.985e-04] | 5.598e-04 | $\begin{pmatrix} 0.1289 \\ 0.6474 \end{pmatrix}$ | 3.756e-04<br>3.740e-04 | [3.586e-04 ; 3.926e-04]<br>[3.567e-04 ; 3.913e-04] |
| 40-49 | 1.030e-03 | [1.004e-03 ; 1.055e-03] | 1.478e-03 | $\begin{pmatrix} 0.06265 \\ 0.5193 \end{pmatrix}$ | 9.583e-04<br>9.468e-04 | [9.165e-04 ; 1.000e-03]<br>[9.034e-04 ; 9.901e-04] |
| 50-59 | 2.828e-03 | [2.774e-03 ; 2.882e-03] | 3.634e-03 | $\begin{pmatrix} 0.03195 \\ 0.458 \end{pmatrix}$ | 2.671e-03<br>2.640e-03 | [2.570e-03 ; 2.772e-03]<br>[2.534e-03 ; 2.746e-03] |
| 60-69 | 7.110e-03 | [7.017e-03 ; 7.202e-03] | 8.178e-03 | $\begin{pmatrix} 0.03663 \\ 0.6993 \end{pmatrix}$ | 6.858e-03<br>6.818e-03 | [6.605e-03 ; 7.112e-03]<br>[6.552e-03 ; 7.083e-03] |
| 70-79 | 0.01776 | [0.01749 ; 0.01803] | 0.02207 | $\begin{pmatrix} 0.1359 \\ 0.7598 \end{pmatrix}$ | 0.01707<br>0.01704 | [0.01616 ; 0.01799]<br>[0.01611 ; 0.01796] |
| 80-89 | 0.05113 | [0.04955 ; 0.0527] | 0.0622 | $\begin{pmatrix} 0.03923 \\ 0.6318 \end{pmatrix}$ | 0.04935<br>0.04896 | [0.04569 ; 0.053]<br>[0.04513 ; 0.05279] |
| 90+ | 0.1563 | [0.153 ; 0.1597] | 0.1682 | 0.9554<sup>†</sup> | 0.1607 | [0.1504 ; 0.171] |

